# Viral loads observed under competing strain dynamics

**DOI:** 10.1101/2021.07.27.21261224

**Authors:** James A. Hay, Lee Kennedy-Shaffer, Michael J. Mina

## Abstract

A plausible mechanism for the increased transmissibility of SARS-CoV-2 variants of concern (VOCs) results from VOC infections causing higher viral loads in infected hosts. However, investigating this hypothesis using routine RT-qPCR testing data is challenging because the population-distribution of viral loads changes depending on the epidemic growth rate; lower cycle threshold (Ct) values for a VOC lineage may simply reflect increasing incidence relative to preexisting lineages. To understand the extent to which viral loads observed under routine surveillance systems reflect viral kinetics or population dynamics, we used a mathematical model of competing strain dynamics and simulated Ct values for variants with different viral kinetics. We found that comparisons of Ct values obtained under random cross-sectional surveillance were highly biased unless samples were obtained at times when the variants had comparable growth rates. Conversely, comparing Ct values from symptom-based testing was largely unaffected by epidemic dynamics, and accounting for the time between symptom onset and sample collection date further reduced the risk of statistical errors. Finally, we show how a single cross-sectional sample of Ct values can be used to jointly estimate differences in viral kinetics and epidemic growth rates between variants. Epidemic dynamics should be accounted for when investigating strain-specific viral kinetics using virologic surveillance data, and findings should be corroborated with longitudinal viral kinetics studies.

## Introduction

One of the biggest threats to combating the SARS-CoV-2 pandemic, or indeed any virologic epidemic, is the emergence of novel variants that may be harder to control or exhibit increased disease severity [1–4]. Variants with increased growth rates, which may arise through mutations affecting infectivity or antigenicity, can quickly come to dominate existing lineages and generate new waves of infection [5–11]. Whereas existing non-pharmaceutical interventions and population immunity may be sufficient to suppress existing viruses, they may be insufficient for more transmissible variants, threatening the robustness of vaccine-induced herd immunity and exacerbating problems in settings with limited access to vaccines. Identifying measurable properties of variants that indicate increased transmission potential is therefore essential for controlling the SARS-CoV-2 pandemic, as in the case of seasonal influenza [12–14].

A hypothesized mechanism for increased transmissibility relates to improved within-host replication, which may result in higher viral loads [15–17]. If viral load predicts infectivity [17,18,19], then infections with new variants that elicit higher peak viral load, shorter incubation periods or slower clearance rates could be more transmissible and for a longer period of time. Testing this hypothesis would ideally rely on longitudinal viral kinetics studies to directly compare viral loads over the course of infection [16,20,21]. However, such data are rare, and comparisons of viral loads have therefore typically been done using routinely collected RT-qPCR surveillance data from asymptomatic or symptomatic individuals [5, 22]. Indeed, multiple such studies have now proposed that samples isolated from variant of concern (VOC) infections demonstrate higher viral loads than from non-VOC infections [22–26].

However, comparisons of viral loads, usually proxied using RT-qPCR cycle threshold (Ct) values, from surveillance samples are potentially biased depending on the epidemiological context [27–29]. It has been shown that cycle threshold (Ct) values observed through population-level surveillance are expected to change depending on the underlying epidemic growth rate: Ct values are skewed lower when the epidemic is growing due to the abundance of recent infections, and skewed higher when the epidemic is growing due to the predominance of older infections [27, 30]. Comparing Ct values from different lineages which may have different growth rates at the time of sample collection therefore has the potential for bias. Higher viral loads from VOC samples may simply reflect a growing epidemic as opposed to higher peak or more sustained viral loads, making it difficult to accurately infer differences in underlying viral kinetics. Methods are needed to accurately quantify the contribution of virologic changes to viral load dynamics at the within-host and population levels.

Here, we explore in simulation different virologic and surveillance scenarios where epidemic dynamics can confound viral load comparisons between variants. We first demonstrate how average viral loads observed at the population level from new, more transmissible variants would be expected to differ from existing viruses even with identical post-infection viral kinetics. We then demonstrate how observations of these patterns differ depending on whether samples are obtained through random cross-sectional surveillance or symptom-based testing. We show that accounting for the epidemic growth rate when comparing samples from random cross-sectional surveillance or time-since-onset when comparing samples from symptom-based surveillance can lead to robust comparisons of RT-qPCR results between variants with different epidemic dynamics. Finally, we present a method for comparing growth rates from different variants using a single cross-section of Ct values whilst accounting for potential differences in underlying viral kinetics.

## Results

### Modeling framework to compare viral loads arising in a two-strain epidemic

We developed a mathematical model to simulate viral loads, observed as RT-qPCR Ct values, in a population undergoing a two-strain epidemic (Figure 1). We implemented a two-strain SEIR model to simulate incidence curves under a scenario where a more transmissible variant (referred to as the “new variant”) is introduced into the population and outcompetes a preexisting, less transmissible lineage (referred to as the “original variant”). We assumed that these strains have identical epidemiological parameters other than their basic reproductive numbers, and that infection elicits symmetric cross-protection against the other variant. We combined these incidence curves (Figure 1A) with the viral kinetics models shown in Figure 1B to simulate viral loads for infected individuals over time. We compared two scenarios for underlying viral kinetics: 1) both variants have identical viral kinetics and 2) the new variant has a higher peak viral load and a slower clearance rate. To simulate observed viral loads, individuals were randomly sampled under one of two strategies: either 1) random cross-sectional surveillance, where individuals are sampled from the population at random regardless of their infection status, or 2) symptom-based surveillance, where individuals are tested after some delay following the onset of symptoms. These scenarios underpin all analyses up to the section “*Quantifying differences in growth rate and viral kinetics of variants using cross-sectional Ct values*” unless stated otherwise.

**Figure 1.**
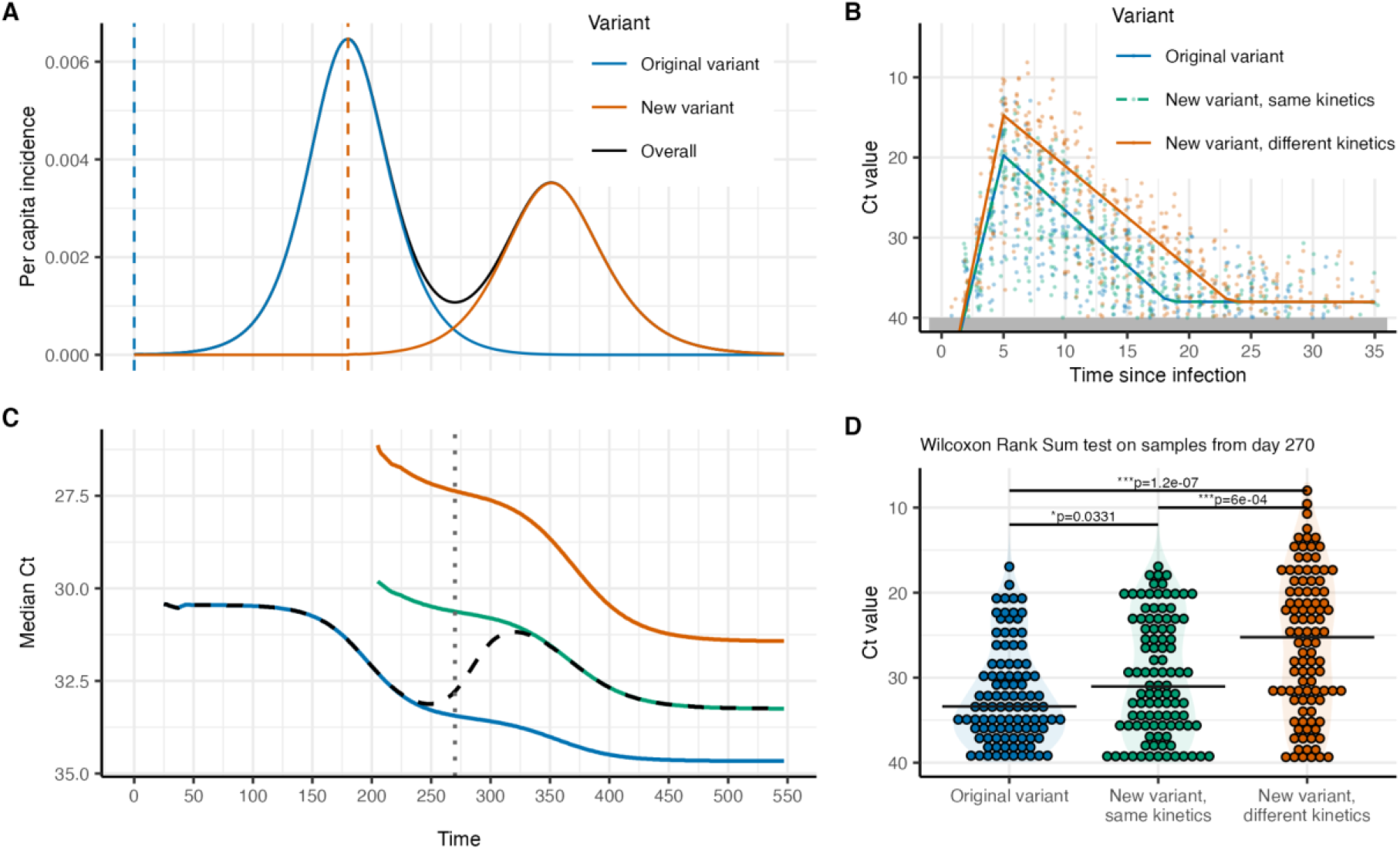
Variants with different epidemic growth rates will exhibit differences in observed average viral loads regardless of true differences in within-host kinetics. (**A**) Incidence curves from a two-strain susceptible-exposed-infected-recovered (SEIR) model, where a virus introduced on day 0 (vertical blue line) with *R_0_*=1.5 is outcompeted by a new variant introduced on day 180 (vertical red line) with *R_0_*=2.5, leading to two waves of infection. (**B**) Variants are assumed to follow either identical or different viral load kinetics, with modal viral loads (solid lines) peaking within one week post infection, coinciding with the typical time of symptom onset, and then declining to near the limit of detection at around three weeks post infection. Substantial variation in individual Ct values (individual points) are observed due to individual-level kinetics and sampling variation. (**C**) The median Ct value observed from individuals sampled entirely at random will reflect the growth rate of that variant at the time of sampling. Black dashed line shows the overall median Ct value. (**D**) Comparison of simulated Ct values obtained on day 270 using a Wilcoxon rank sum test. When the original variant is in decline and the new variant is growing, the comparison reflects a significant difference in viral load between the variants regardless of a true difference in underlying viral kinetics.

### Comparing viral loads from samples with different growth rates

The simulations show that the distribution of viral loads among infected individuals changes over time, reflecting the growth rate of the epidemic (Figure 1C). Because the two variants have different transmission rates and introduction dates, their viral load distributions differ at any given point in time regardless of any true difference in viral kinetics. Under random cross-sectional surveillance, this difference arises because the time-since-infection distribution changes over the epidemic: randomly sampled infections are typically more recent when the epidemic is growing than when it is declining [27, 30]. As a result, statistical tests comparing Ct values from samples obtained under random cross-sectional surveillance at a single point in time will reflect differences arising both from epidemic dynamics, which dictate the recency of infection, as well as underlying viral kinetics (Figure 1D). When simulated samples were obtained in this way and the new variant samples were compared to the original variant samples using a Wilcoxon rank-sum test with significance level of 5%, we found that type 1 statistical errors occurred in 25% of simulations when using 25 detectable Ct values from each of the two variants, increasing to nearly 100% when comparing 500 detectable Ct values (Figure S1).

One approach to overcome these statistical biases and accurately detect underlying differences in viral kinetics is to compare samples taken from time points of comparable growth rates (Figure 2A). For example, comparing original variant samples taken during the first wave of infections to new variant samples taken during the second wave will lead to more accurate statistical tests for underlying differences in viral kinetics (Figure 2B). Comparing samples in this way resulted in type 1 errors in only 5% of trials (the nominal rate), with statistical power of at least 95% when 250 or more detectable Ct values per variant were sampled (Figure S2&S3).

**Figure 2.**
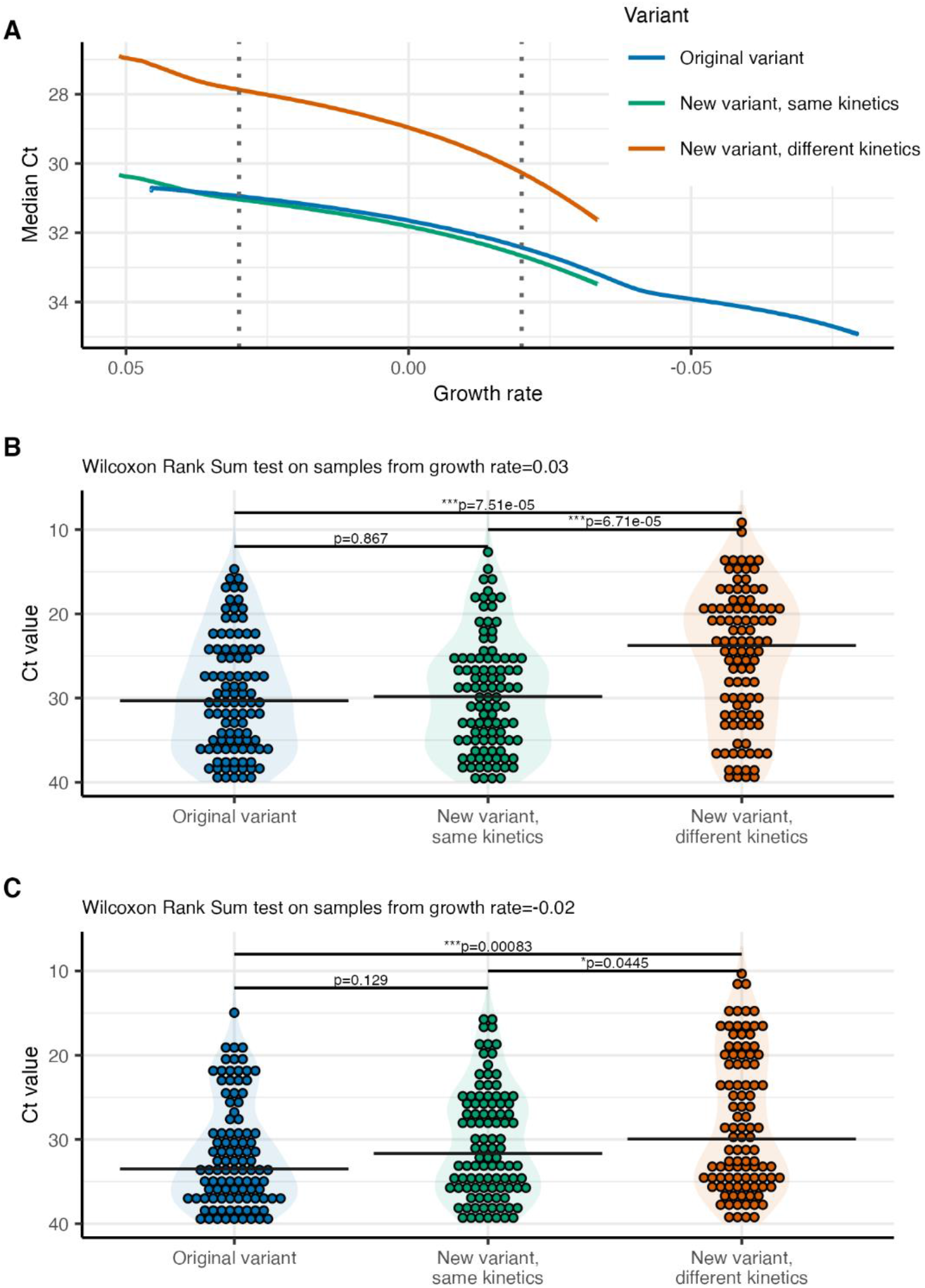
Comparing Ct values obtained from variants at times of comparable epidemic growth lead to accurate comparisons of viral loads. (**A**) Using the simulations shown in Figure 1C, median Ct values for the variants are plotted against their growth rate at time of sampling. (**B**) Comparison of simulated Ct values obtained from the time when each variant had a growth rate of 0.03 (i.e., the log ratio of new infections tomorrow relative to today) using a Wilcoxon Rank Sum test. (**C**) As in (B), but when the growth rate is -0.02.

### Viral loads from symptom-based surveillance more accurately reflect underlying differences in viral kinetics

In reality, most RT-qPCR results are obtained from non-random sampling. In particular, symptom-based surveillance, where individuals with recent symptom onset seek testing, is likely to comprise the majority of samples. To test whether epidemiological biases were present in Ct values obtained under symptom-based surveillance, we simulated Ct values using the same time-since-infection model in Figure 1C but assumed that individuals were sampled after some delay following the onset of symptoms. We assumed that symptomatic individuals have incubation periods drawn from a log-normal distribution with median 5.0 days and variance of 5.8 days, and are subsequently sampled after some delay drawn from a discretized gamma distribution with shape and scale parameters of 5 and 1 respectively (Figure 3A). This corresponds to a mean confirmation delay of 4.5 days and variance of 5.1 days following symptom onset. Both variants were assumed to have the same incubation period and sampling delay distribution. In addition, on the individual level, the Ct value at any day was independent of the incubation period length and sampling delay. We note that the choice of these distribution parameters is arbitrary; similar patterns will be observed with different values.

**Figure 3.**
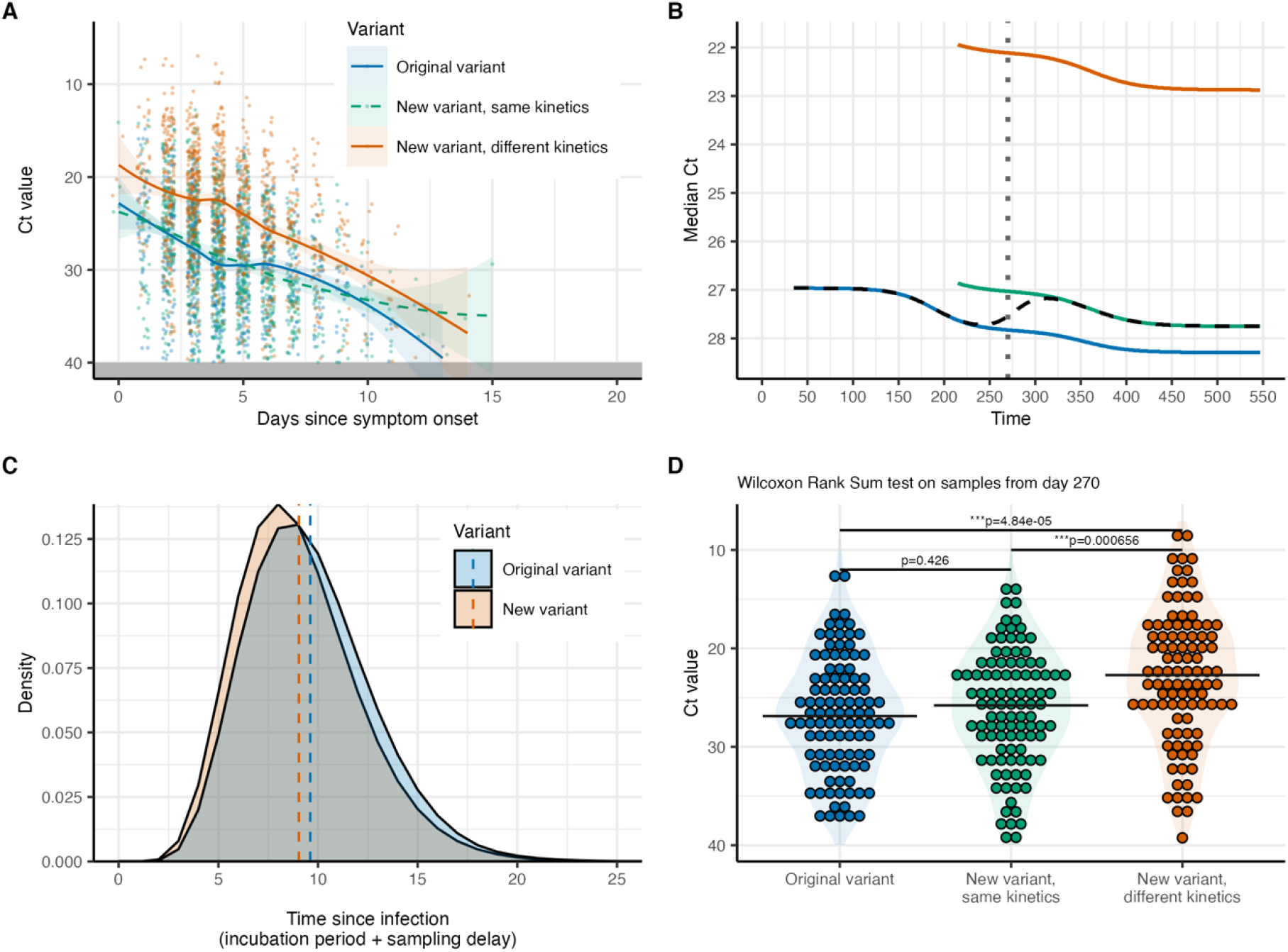
Ct values obtained through symptom-based surveillance more accurately reflect true viral load differences. (**A**) Ct values simulated under the same time-since-infection model as in Figure 1, but with samples taken after the onset of symptoms and some additional sampling delay. Time from infection to symptom onset was drawn from a log-normal distribution with median 5.0 days and variance 5.8 days. Time from symptom onset to sample collection was drawn from a discretized gamma distribution with mean 4.5 days and variance 5.1 days. Solid line and ribbons show fitted smoothing spline and 95% CI. (**B**) Median Ct value observed over time from individuals sampled under symptom-based surveillance. (**C**) Distribution of incubation periods for individuals sampled on day 270 of the simulation, stratified by variant. Note that the observed distribution on day 270 differs from the log-normal distribution used for simulation through its convolution with the infection incidence curve. (**D**) Comparison of simulated Ct values obtained on day 270 under symptom-based surveillance using a Wilcoxon rank sum test.

Although some differences remained in observed Ct values over time between the two variants, the difference was small unless the new variant had truly different underlying viral kinetics (Figure 3B). This is because the time-since-infection distributions for the two strains are comparable regardless of the underlying growth rate, as individuals are always sampled at a similar time post onset and therefore post infection (Figure 3C). Therefore, comparing Ct values between variants obtained from symptomatic surveillance largely reflects true differences in underlying viral kinetics (Figure 3D).

However, statistical comparisons may still lead to flawed conclusions if the time-since-onset and sampling delay distributions are substantially different between the two variants. Even when the same distributions are assumed, some small difference in the observed time-since-infection distribution will arise due to differences in the underlying epidemic growth rate (see Figure S3 and S4 from [27] for further details). For example, if an epidemic is growing, then individuals with symptom onset on a given day are more likely to have been recently infected with a short incubation period simply due to the abundance of recent infections. Our simulations show that as the number of samples being compared increases, the probability of encountering a type 1 statistical error when using the Wilcoxon rank sum test increases. This is true because of the reduced variation within groups, even though the between-variant estimated median Ct difference is the same (Figure S4).

Using a linear regression model and including days-since-onset as an explanatory variable drastically reduces the frequency of type 1 errors (Figure S5). Indeed, accounting for days-since-onset becomes vital when the distribution of delays between symptom onset and sampling differs between the two variants. Figure S6 shows type 1 statistical errors are almost guaranteed when using a Wilcoxon rank sum test if the original variant has a different sampling delay distribution (assuming shape and scale parameters of 7 and 0.9 as opposed to 5 and 1), but Figure S7 shows that these errors can be drastically reduced through using a regression model accounting for days-since-onset. We note that type 1 errors still arise at large sample sizes, as accounting for days-since-onset only partially accounts for the small differences in the time-since-infection distribution (i.e., it does not account for the incubation period).

### Quantifying differences in growth rate and viral kinetics of variants using cross-sectional Ct values

Finally, we approached the system from a different perspective: rather than testing for differences in viral kinetics between variants based on surveillance samples, we asked if we could use random cross-sectional samples taken from the same point in time to infer differences in variant-specific growth rates. Again, we generated a synthetic population of SARS-CoV-2 infections by simulating Ct values obtained through random cross-sectional surveillance at a single point in time assuming that the variants had different viral kinetics (as shown in Figure 1B). We then adapted a previously described method to estimate the growth rate and viral kinetics parameters based on single cross-sections of Ct values [27]. We tested two epidemiologic scenarios: 1) incidence of the original variant declines while the new variant increases and 2) both variants have positive growth rates at the same time, but the new variant has a higher basic reproductive number (*R_0_*=2.5 vs. 1.5).

When 100 detectable Ct values for each variant were obtained through random cross-sectional surveillance at a time when one variant was declining and the other was increasing in frequency (Figure 4A), we were generally able to accurately re-estimate the true growth rates of the two variants (Figure 4B). The model was also able to quantify the true difference in peak viral loads elicited by the new variant (Figure 4C); however, it was not able to identify the slower clearance rate. Similarly, in the scenario when infections from both variants were simultaneously increasing but at different rates (Figure 4D), we were able to identify that the new variant likely had a higher growth rate, although not unequivocally based on the 95% credible intervals (CrI) on the difference in growth rate (Figure 4E). Again, differences in peak Ct value were identifiable, but not in the clearance rate (Figure 4F). These results show that although lower median Ct values may be explained either by higher growth rates or differences in viral kinetics, the full distribution of Ct values may hold information that allows both processes to be identified.

**Figure 4.**
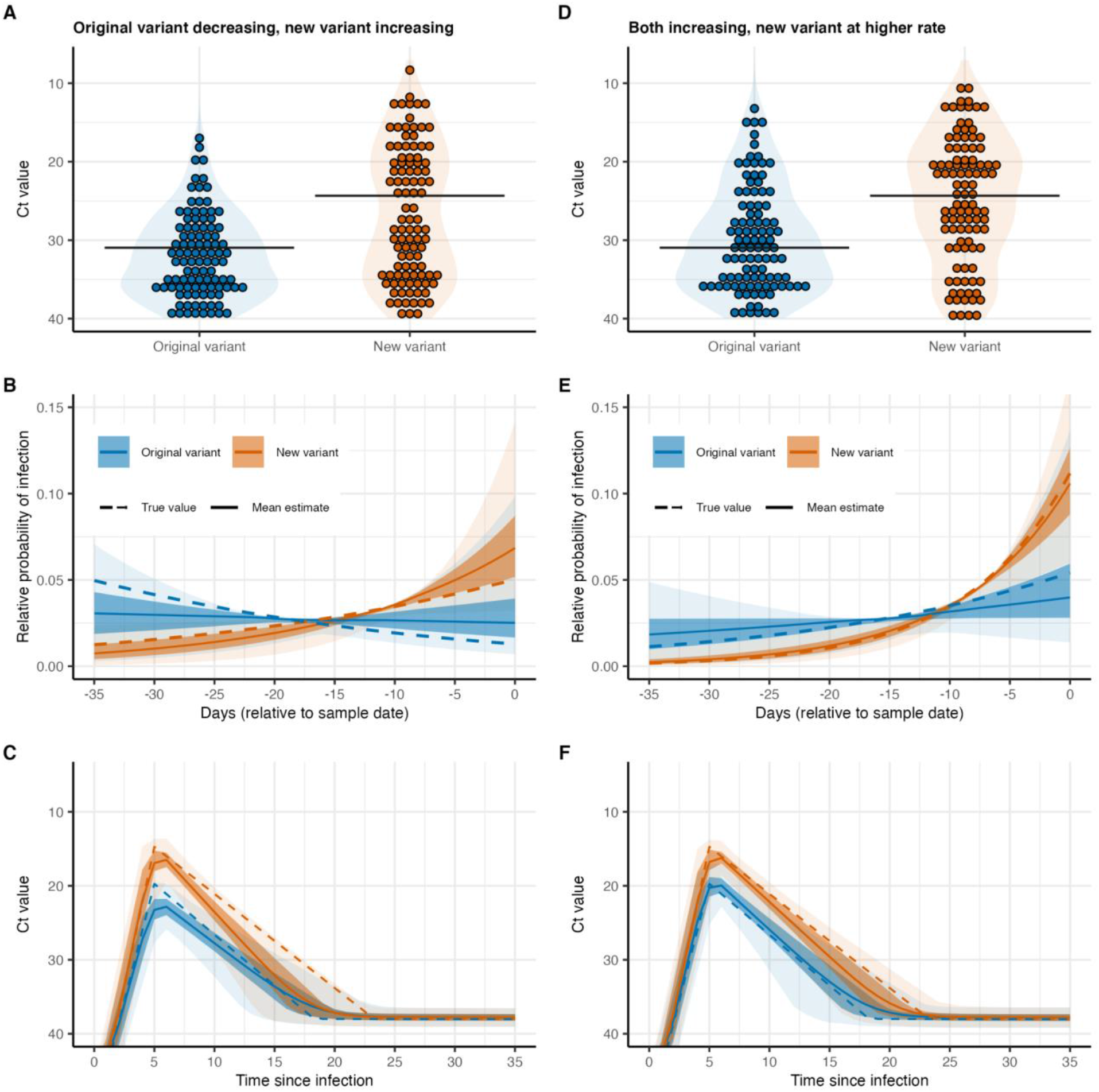
(**A**) Simulated Ct values assuming that the new variant has a higher peak viral load (−5 Ct at peak) and a longer time to clearance (5 additional days from peak to second hinge). Samples were assumed to be obtained under random cross-sectional surveillance at day 270 of the simulation shown in Figure 1. Horizontal lines show median Ct. (**B**) Estimated epidemic trajectory inferred using the single cross section of Ct values. Solid line, dark shaded region and light shaded region show posterior mean, 50% credible intervals (CrI) and 95% CrI respectively. Dashed line shows the true epidemic trajectory for each variant. (**C**) Inferred viral kinetics from the same model fitting procedure as in (B). Shown are the posterior estimates for the modal Ct value over time since infection. Solid line, dark shaded region and light shaded region show posterior mean, 50% credible intervals (CrI) and 95% CrI respectively. Dashed line shows the true modal Ct curve. (**D**), (**E**), and (**F**) are identical to (A), (B) and (C) respectively, but with Ct values obtained from a simulation where both variants are seeded at the same time, but with the new variant assumed to have a higher basic reproductive number.

The results shown in Figure 4 are from a single simulation of detectable Ct values. Therefore, to explore the identifiability of these differences in growth rates and viral kinetics based on cross-sectional samples of different sizes and stochastic draws, we repeated the analyses with varying sample sizes from 25 to 500 detectable Ct values per variant, running 100 simulations for each sample size. When one variant was in decline while the other was increasing in frequency (Figure S8), we generally identified the different direction of the epidemic trajectories with as few as 25 Ct values per variant (with increasing certainty at increasing sample sizes). However, when both variants were increasing at the same time but at different rates (Figure S9), we were not able to exclude the possibility of no difference in growth rate even with 500 Ct values per variant. This appeared to be largely driven by the lack of precision in growth rate estimates for the original variant, resulting in wide credible intervals. Nonetheless, posterior mean estimates were consistently identified as different between the two variants. Differences in peak Ct value were also identifiable in both epidemiologic scenarios with as few as 25 Ct values; however, even at 500 Ct values per variant, we were unable to reliably identify the true difference in viral clearance rates (Figures S10 and S11).

## Discussion

Given the relationship between epidemic growth rate and the distribution of viral loads in a population [27], care must be taken when directly comparing viral loads between variants using surveillance samples to avoid drawing incorrect conclusions of differences in strain virulence. Infections from variants with positive growth rates will typically be more recent than infections from co-circulating variants with negative growth rates, resulting in higher average viral loads. These findings provide a conceptually similar caution to analyses assessing the association between mutations and transmissibility [14, 31]: just as mutations may be associated with increased growth rates simply due to founder effects or chance, infections with new variants may be associated with increased viral loads simply because they are experiencing higher epidemic growth rates.

We found that comparing Ct values between variants using samples obtained entirely at random from the population are far more likely to lead to incorrect conclusions regarding differences in underlying viral kinetics than samples obtained from recently symptomatic individuals. This is because of how these sampling strategies reflect the underlying time-since-infection distribution: cross-sectional surveillance samples individuals at a random time point in their infection, whereas symptom-based surveillance systematically tests people at a similar time after infection. Because viral loads [32], and therefore Ct values, are dictated by the time post infection, comparing Ct values from samples with similar time-since-infection distributions will reflect differences in viral kinetics and not epidemic growth rates. Accounting for differences in the time-since-infection distribution between datasets, which may be achieved using samples taken from times of comparable growth rates or by including time-since-onset or time-since-infection in a regression model, will improve the reliability of statistical tests comparing Ct values between variants. However, these results assume that the variants have similar delays between exposure and symptom onset. If, for example, newer variants have systematically shorter symptom onset delays, then new variant samples obtained under symptom-based surveillance would reflect a shorter time-since-infection distribution.

At the time of writing, the SARS-CoV-2 literature has a mixture of studies that either do not acknowledge the potential for biases in Ct value comparisons [6, 26], discuss the potential for this bias [33, 34], or take clear steps to account for differences in the time-since-onset or time-since-infection distribution when comparing Ct values [5, 22]. For example, an analysis of P.1 samples (Gamma variant) in Manaus, Brazil found that Ct values declined over time as the prevalence of P.1 infections increased. An initial comparison of values found a statistically significant association between P.1 infection and lower Ct value; however, after accounting for the delay between symptom onset and sample collection, the significance of this relationship was lost, similar to our simulation results [5]. Another analysis comparing Ct values between two variants in Washington State, USA also initially performed a direct comparison of Ct values between a variant with a 614G substitution [22]. Again, the authors found that 614G was associated with lower Ct values than 614D. In contrast to the analysis by [5], this significant difference in Ct value remained after accounting for a number of potential confounders such as days after symptom onset and patient age. In this study, epidemic dynamics were likely not important in dictating Ct value distributions, as Ct values did not appear to be associated with time of sample collection and the time-since-onset distribution was similar between the two variants.

Although epidemic dynamics have the potential to bias viral load comparisons, particularly using random cross-sectional samples, many study designs are unlikely to be affected. Comparing samples from recently symptomatic individuals, particularly when the distribution of delays between symptom onset and sample collection date are similar between the variants or are included in a regression model, is likely to lead to reliable conclusions. Such comparisons will be even more reliable if the timing of symptom onset is dependent on viral load (i.e., symptom onset occurs as a result of viral loads reaching their peak as opposed to independently). It is worth noting that differences in sampling delay distributions will likely vary independently of the epidemic growth rate simply due to the logistics of sample collection, limitations on testing capacity and changes in policy dictating who is tested. Accounting for time-since-onset is therefore advisable regardless. Other instances where epidemic dynamics will not affect viral load comparisons are when samples are obtained longitudinally from the same individuals, allowing the comparison of the full viral kinetics curve [20, 35], or obtained near the time of exposure such that all samples have a similar time-since-infection [36].

There are a number of additional factors affecting viral load distributions that we did not consider here. First, we assumed in our simulations that Ct values were comparable across all samples. In reality, Ct values from different lineages may not be comparable across primers and platforms, such that Ct value comparisons reflect technological limitations rather than differences in viral load [37]. In such cases, conversion to a common scale such as viral load using calibration curves may be advisable [17]. We also did not consider how patient-level factors affect viral loads. Some limited evidence exists to suggest that children exhibit systematically lower viral loads than adults, with observations also affected by external factors such as swab quality or viral location within the respiratory tract [16,38,39]. Clinical severity may also affect viral load kinetics, with symptomatic patients exhibiting slower clearance rates than asymptomatic infections [32, 40]. If samples being compared between variants represent different underlying populations, which is likely when the age distribution of cases shifts over time, then viral load differences may reflect differences in the tested population rather than viral kinetics. Such shifts in clinical severity and age distributions have been seen in countries with high vaccine coverage, as vaccination of the elderly and at-risk groups shifts the relative prevalence of infections into younger populations [41]. Additionally, vaccination not only affects the composition of the population testing positive for SARS-CoV-2, but infections in vaccinated individuals may also exhibit lower viral loads [42]. Overall, care should be taken to either compare sample sets from similar populations, or meta-data on relevant demographic factors should be accounted for when comparing Ct values between variants.

Finally, we adapted a previous method, which is ultimately a generalized linear regression model, to use Ct values obtained from a single point in time to simultaneously estimate differences in growth rates alongside differences in viral kinetics [27]. In reality, these two mechanisms may not be uniquely identifiable; however, understanding the combination of viral kinetics and transmissibility differences that can explain the data may still be valuable to detect variants of concern early and to quantify transmission advantages. This approach may be particularly useful in settings where sequencing capacity is limited, as sequencing-independent means of stratifying samples by lineage (e.g., variant-specific primers, single gene failure etc.) can be used to then compare Ct values between variants [43]. However, we emphasize that this approach currently requires samples obtained through random cross-sectional surveillance, or nearly random samples such as non-COVID patient hospital testing.

## Materials and methods

### SEIR model

We used a deterministic, two-strain, susceptible-exposed-infected-recovered (SEIR) model to simulate incidence curves for two competing strains in a fully susceptible population [44]. We assumed that two strains were seeded in the population: an “original variant” with *R*_0_ = 1.5, and a “new variant” with *R*_0_= 2.5. The “original variant” was seeded on day 0 of the simulation, and the “new variant” was either seeded on day 180 or on day 0 as specified in the *Results*. We assumed that both strains had a 3-day mean latent period and 7-day mean infectious period, and that individuals became permanently immune to the strain they were infected with

upon recovery. We also assumed that infection with one strain elicited strong cross-immunity (75% reduced infection probability) to the other. Overall, this is a simplified implementation of the model presented by [44] with no seasonality, no waning immunity and no births or deaths. The model was solved using daily time steps, and daily growth rates were calculated as *g*(*t*) = 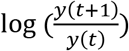 where *y*(*t*) is the incidence of new infections on day *t*. Full model equations and a table of parameters are shown in the *Supplementary Material: SEIR model equations* and Table S1.

### Viral kinetics model

We used an existing model describing the average and distribution of viral loads as a function of time-since-infection with SARS-CoV-2 [27]. Briefly, the model assumes that the modal Ct value follows a piecewise linear function following infection, *C_mode_*(*a*). After an initial 1-day period of no viral growth, the modal Ct value decreases monotonically to a peak value at day 5 post infection. We assumed the peak Ct value was 20 for the original variant, or 15 for the new variant in scenarios assuming different viral kinetics. Ct values then increase to a plateau at a value of 38, which occurred on day 13 post peak for the original variant and day 18 post peak for the new variant with different viral kinetics. Thereafter, individuals have a daily probability of becoming fully undetectable, modeled as a Bernoulli process with probability *p_addl_*.

To capture the substantial variation across individuals and samples in Ct values observed on a given day post-infection, we assumed that Ct values follow a Gumbel distribution with a scale parameter *σ*(*a*) that begins at 5 but decreases to 4 towards the end of infection. This decreasing scale parameter captures the fact that samples taken early in infection are affected by variation from both individual-level heterogeneity in kinetics in addition to sampling variation, whereas samples taken towards the end of infection largely represent consistent, small quantities of residual viral RNA. We assumed that the maximum Ct value was 40, and all samples from undetectable or uninfected individuals were excluded from the analyses. We also note that although we describe the model on the scale of Ct values, analogous results would hold for viral loads, as Ct values are linearly related to log viral loads. Full model equations and a table of parameters are shown in the *Supplementary Material: viral kinetics* model and Table S2.

### Simulated surveillance samples and Ct values

We combined the SEIR and viral kinetics models to simulate Ct values among the infected population over time. In brief, we used the SEIR model to simulate when individuals are infected with either variant over time and used the viral kinetics model to simulate their observed Ct value when sampled on a particular day post infection. Observations were simulated using two surveillance strategies described below. For the analyses underpinning Figures S1-S7, we generated 1000 simulated datasets for each sample size of 25, 50, 100, 250 or 500. We simulated datasets on day 270 of the epidemic in scenarios where the new variant had a later seed date, or day 50 when the strains were seeded on the same day.

First, we considered random cross-sectional sampling, where individuals are tested entirely at random regardless of their time since infection. In this case, we simulated the distribution of detectable Ct values *X_v,t_* for variant *v* on a given day of the epidemic *t* as *X_ν,t_*∼*f_ν,t_*. *f_ν,t_*(*x*) is the probability density function (PDF) for *detectable* Ct values on day *t* of the epidemic, calculated by convoluting the variant-specific incidence curve (which describes the distribution of times since infection) and viral kinetics model (which describes the distribution of observed Ct values on each day after infection):

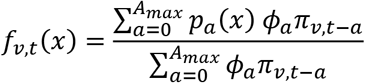

where *A_max_* is the maximum time-since-infection for which individuals may still be PCR detectable (set to 35 days); *p_a_*(*x*) is the Gumbel probability density function scaled to only take values between 0 and *C_LOD_* with location parameter *C_mode_*(*a*) and scale parameter *σ*(*a*); *ϕ*_a_ is the probability of being PCR detectable on day *a* post infection; and *π_v,t_* gives the probability of infection with variant *v* on day *t* of the epidemic. Individual observations were generated by simulating from this PDF. Note that this PDF can be modified to generate PCR-negative observations, but we are only interested in the distribution of PCR-positive observations here. Second, we simulated Ct values under symptom-based surveillance, where individuals are tested following the onset of symptoms. Again, we simulated from the PDF for Ct values on day *t* of the epidemic as *X_ν,t_*∼*g_ν,t_*(*x*), but this time convoluting the incubation period and sampling delay distributions as well as the incidence and viral kinetics models. Note again that we assume that only PCR-positive Ct values were observed:

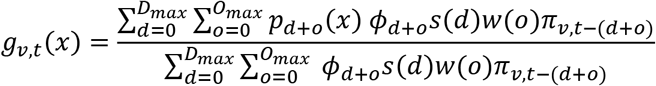

where *d* is the sampling delay in days; *o* is the incubation period in days; *s*(*d*) is the PDF for the discretized gamma sampling delay distribution; and *w*(*o*) is the PDF for the discretized log-normal incubation period distribution.

Simulating Ct values for the analyses shown in Figure S5 and Figure S7 is slightly more involved because each individual needs both an observed Ct value and corresponding sampling delay. Although the principle of simulating from the PDF is the same, we simulated individual-level line list data for these analyses as described in the *Supplementary Material: simulating samples under symptom-based surveillance*.

### Statistical methods

Direct statistical comparisons of Ct distributions from the two variants were two-sided, two-sample Wilcoxon rank sum tests (Mann-Whitney test) at a significance level of 5%. In the analyses controlling for days-since-onset when comparing Ct values, we fit linear regression models of the form *E*[*x_i_*|*d_i_*,*ν*_*i*] = *β*_0_ + *β*_1_*d_i_* + *β*_2_*ν*_i_, where *d_i_* gives the days between symptom onset and sample collection for individual *i*, and *ν_i_* gives the variant infecting that individual. Hypothesis tests in the regression models were conducted for the null hypothesis *H*_0_:*β*_2_ = 0 using an asymptotic t-test. Power was defined as the proportion of simulations under a given scenario and sample size where the null hypothesis (the distribution of Ct values for both variants are equal or there is no effect of variant on Ct value) was correctly rejected in simulations where the variants have different viral kinetics. Type 1 errors were defined as tests which incorrectly rejected the null hypothesis in simulations where the variants have identical viral kinetics.

### Estimating growth rates and viral kinetics from cross-sectional samples

Using the simulated random cross-sectional samples of detectable Ct values, we jointly estimated the posterior distributions of viral kinetics parameters and the exponential growth rate that are consistent with the observed data. We repeated this process for each of the 100 simulated datasets of sample sizes of 25, 50, 100, 250 and 500 detectable Ct values per variant. Model fitting was carried out using the *virosolver* R package [45]. In brief, model fitting involves: (1) defining the likelihood of observing Ct values conditional on the viral kinetics model parameters and incidence curve, assuming that incidence follows exponential growth with an unknown growth rate parameter; (2) defining priors for all model parameters as described in Table S2; and (3) estimating the posterior distribution of all model parameters conditional on the Ct data using Markov chain Monte Carlo (MCMC). For each model fit, three chains were run for 100,000 iterations each with the first 50,000 iterations discarded as burn in. Convergence was assessed based on the trace plots and obtaining effective sample sizes >200 and *R* < 1.1 for all estimated parameters.

For the most part, we followed the method exactly as described in [27]. However, because we are interested in jointly estimating the viral kinetics and epidemic growth rates for two co-circulating strains (whereas [27] is defined for only one strain), we made the following modifications: (1) the peak Ct value parameter for the new variant was defined relative to the peak Ct value for the original variant (i.e., *C*’_p_= ρ*C_p_*); (2) the second hinge point of the Ct model for the new variant was defined relative to the original variant (i.e., *t*’*_s_* = *ηt_s_*); and (3) each variant has its own exponential growth rate parameter (*β_ν_*).

## Data Availability

All code to perform the analyses and generate the figures presented in this article are available under the GNU General Public License version 3 at https://github.com/jameshay218/variant_viral_loads.

## Acknowledgements

This work was supported by the US National Institutes of Health Director’s Early Independence Award DP5-OD028145 (MJM and JAH).

## Competing interests

MJM is an advisor for Detect, LivePerson and COVID Signals. JAH and LKS declare no competing interests.

## 1. SEIR model equations

Shown below are the ordinary differential equations for the two-strain susceptible-exposed-infected-recovered model. Each equation shows the transition rate for a given state pair. For example, {*S*_1_,*S*_2_} gives the population who are susceptible to both strain 1 and 2, {*S*_1_,*S*_2_} gives the population who are susceptible to strain 1 and exposed to strain 2 etc. For concision, all individuals infected with a given strain are denoted ***I_ν_***, where ***I***_1_ = {*I*_1_*S*_2_}+{*I*_1_*E*_2_}+{*I*_1_*I*_2_}+{*I*_1_*R*_2_} and ***I***_2_ = {*S*_1_*I*_2_}+{*E*_1_*I*_2_}+{*I*_1_*I*_2_}+{*R*_1_*I*_2_}. Note that epidemic seeding is also included here, where |*seed* < *t* ≤ *seed* + 7|k indicates that exposed individuals are generated at rate *κ* within a 7-day window following seeding. Parameters are described in Table S1. Note that 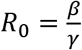.

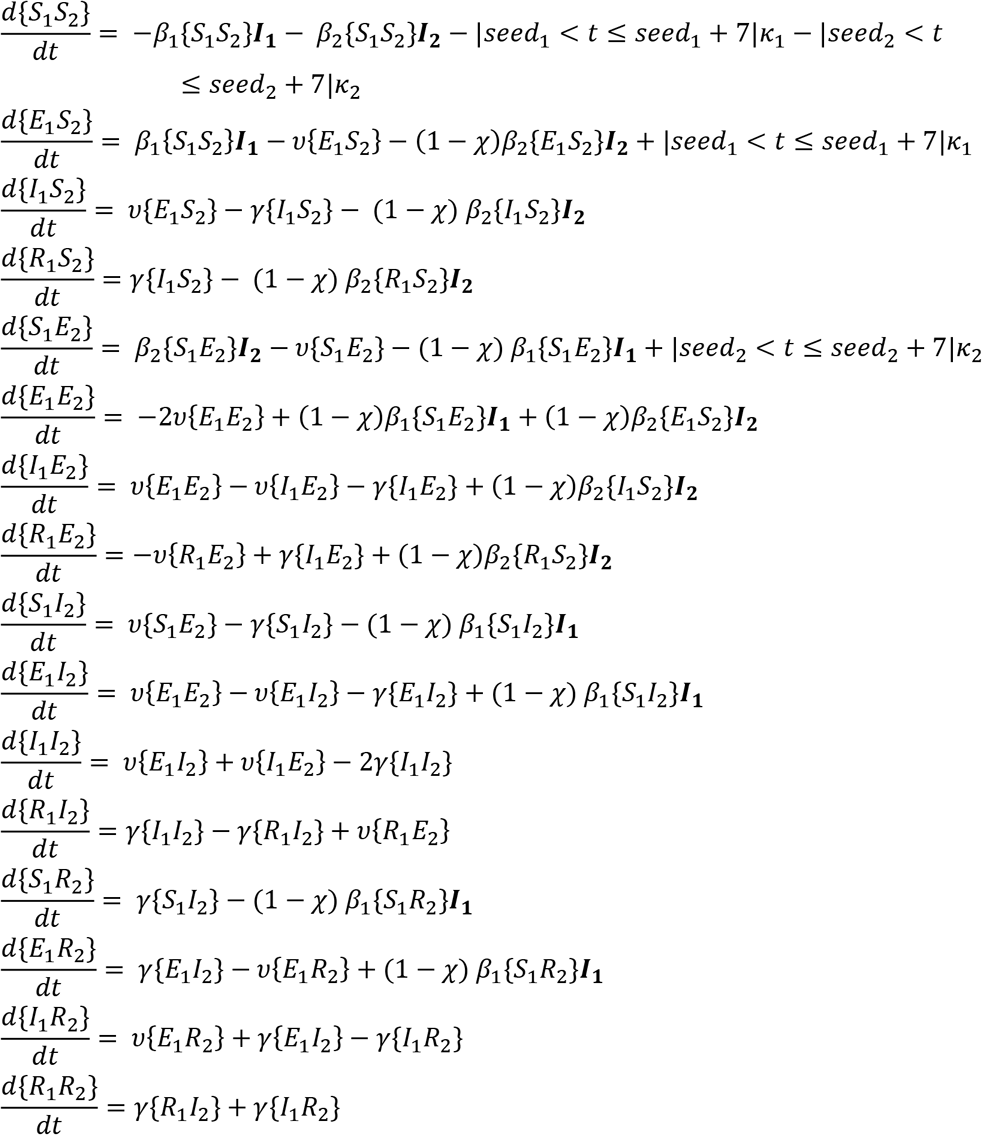

## 2. Viral kinetics model

To describe modal Ct values *C_mode_*(*a*) in an individual on each day post infection *a*, we used a previously described viral kinetics model (main text reference [27]). We reproduce the equations here for completeness. Overall, the model is a two-hinge function that describes the timing and Ct value of several switch points in the viral kinetics process. After a short period *t_e_* of viral loads remaining unchanged at *C*_0_, the modal Ct value decreases to a minimum of *C_p_* over a period of *t_p_* days. Ct values then increase over *t_s_* days to plateau at *C_s_*. From this time onward the distribution of Ct values among detectable individuals remains unchanged; however, to account for individuals fully recovering and becoming PCR negative, we modeled a Bernoulli process with a daily probability *p_addl_* of becoming undetectable on each day after the final hinge point (*a* < *t_e_*+*t_p_*+*t_s_*).

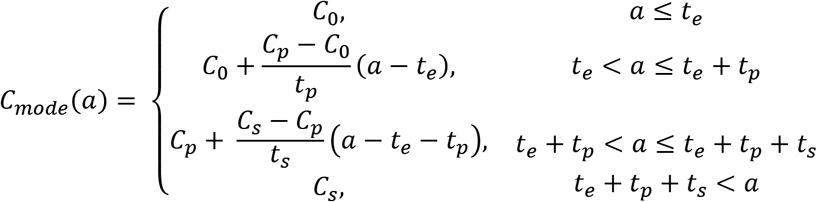

To capture variation in observed Ct values arising from sampling variation and individual-level variation in viral kinetics, we assumed that observed Ct values follow a Gumbel distribution with location parameter set by the above model: *C*(*a*)∼*Gumbel*(*C_mode_*(*a*),*σ*(*a*)). The scale parameter σ(*a*) was assumed to shrink over time given by:

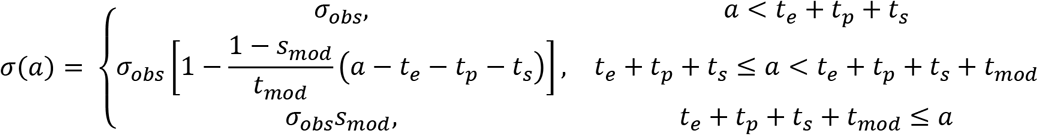

A key part of the model is the description of the proportion of individuals who remain PCR positive on each day post infection. Individual samples can be undetectable in one of two ways: (1) having a Ct value drawn from the Gumbel distribution above the limit of detection *C_LOD_* or (2) fully clearing the infection following the Bernoulli process described above. The probability of being detectable on day *a* post infection is therefore given by:

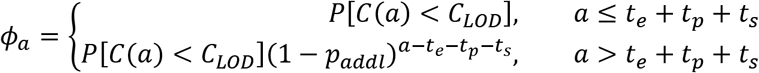

## 3. Simulating samples under symptom-based surveillance

To generate complete line lists for the simulated data underpinning Figure S5 and Figure S7, we used the SEIR model, viral kinetics model, incubation period distribution and sampling delay distribution to generate a population of size *N* where each individual has a binary infection state, an infection time, a binary symptomatic state, an incubation period, a sampling delay, and an observed Ct value. All time-related variables are in days. Note that the incidence curve is generated only by the SEIR model before individual symptom onset dates are simulated—individual symptomatic states do not impact transmission dynamics.

First, we generated an infection state (1 or 0) for each variant for each individual from a Bernoulli distribution with probability 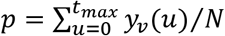 where *y_ν_*(*t*) is the incidence of new infections with variant *v* at time *t*. We then simulated an infection time for each infected individual with each variant *t_ν,i_*:

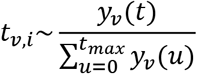

Next, we generated a symptomatic state for each infected individual from a Bernoulli distribution with probability *p* = 0.35. Symptomatic individuals then have an incubation period drawn from:

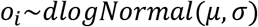

Where *dlogNormal* is the discretized log-normal distribution with mean *μ* and standard deviation *σ*. We set *μ* = 1.621 and *σ* = 0.418 based on previous estimates (main text reference [46]). Individuals then have a sampling delay drawn from:

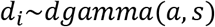

Where *dgamma* is the discretized gamma distribution with shape parameter *a* and scale parameter *s*. We set *a*=5 or 7 and *s*=1 or 0.9 as described in the main text. Infected individuals have a recovery time after which they are guaranteed to be PCR negative, drawn from a negative binomial distribution:

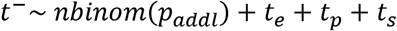

Where *nbinom* is the negative binomial distribution, and the other parameters are described in Table S2. Finally, we simulate an observed Ct value for each individual *i* observed on day *t_ν,i_* + *o_i_* + *d_i_* under the model:

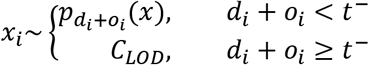

Where 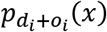 is the Gumbel probability density function with location parameter *C_mode_*(*d_i_* + *o_i_*) and scale parameter *σ*(*d_i_* + *o_i_*. All *x_i_* > *C_LOD_* are set to *C_LOD_*.

To compare Ct values on a given day of the simulation *t_samp_*, we found all individuals in the line list with detectable Ct values where *t_samp_* – 3.5 < *t_ν,i_* + *o_i_*+*d_i_* ≤ *t_samp_* +3.5 (i.e., all individuals who were sampled in a 7 day window around the chosen time), and then resampled from these Ct values with replacement to obtain a sample of the specified size (25, 50, 100, 250 or 500).

---

**Figure S1.**
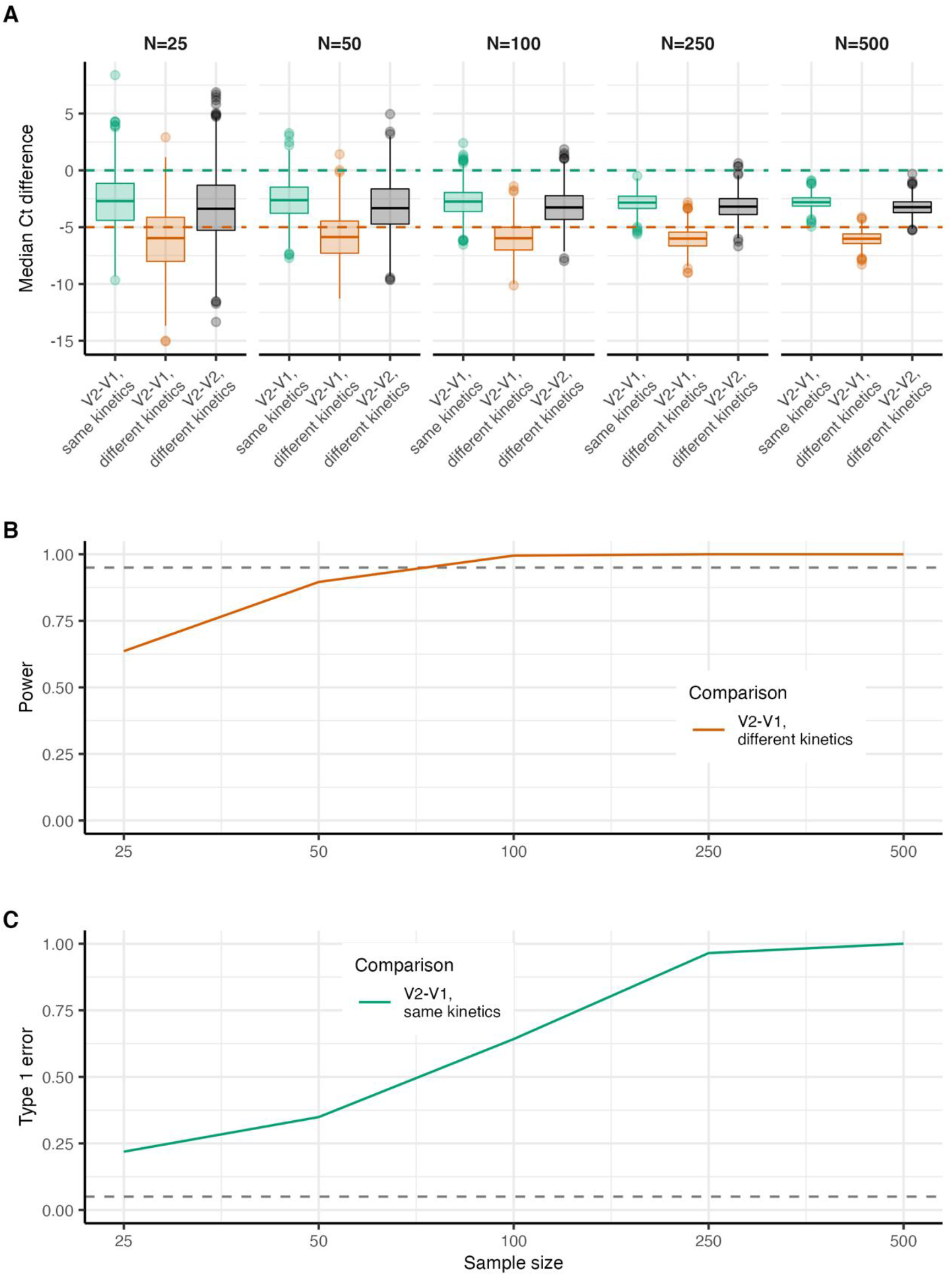
Accuracy and power when comparing Ct values from variants obtained through random cross-sectional surveillance at day 270 of the simulation shown in Figure 1. All statistical tests are two-sided Wilcoxon rank sum tests. (**A**) Boxplots show the interquartile range (IQR, 75th to 25th percentile) and median across 1000 simulations for the difference in Ct values when comparing the original variant (V1), the new variant (V2) with identical kinetics, or the new variant (V2) with different kinetics. Whiskers show 1.5 times the largest and smallest values within 1.5 times the IQR. Dots show individual simulations outside 1.5 times the IQR. Horizontal dashed green lines show the true difference in median Ct values between the original variant and the new variant with identical kinetics. Horizontal dashed orange line shows the difference in peak Ct value assumed for the new variant with different kinetics relative to the original variant. For an entirely accurate test, the green dots should all align on the green horizontal line, and the orange and black dots should be identical and negative. (**B**) Assessment of empirical power to detect a true difference in Ct values at different sample sizes. (**C**) Probability of type 1 error (incorrectly infer a difference in Ct value) at different sample sizes.

**Figure S2.**
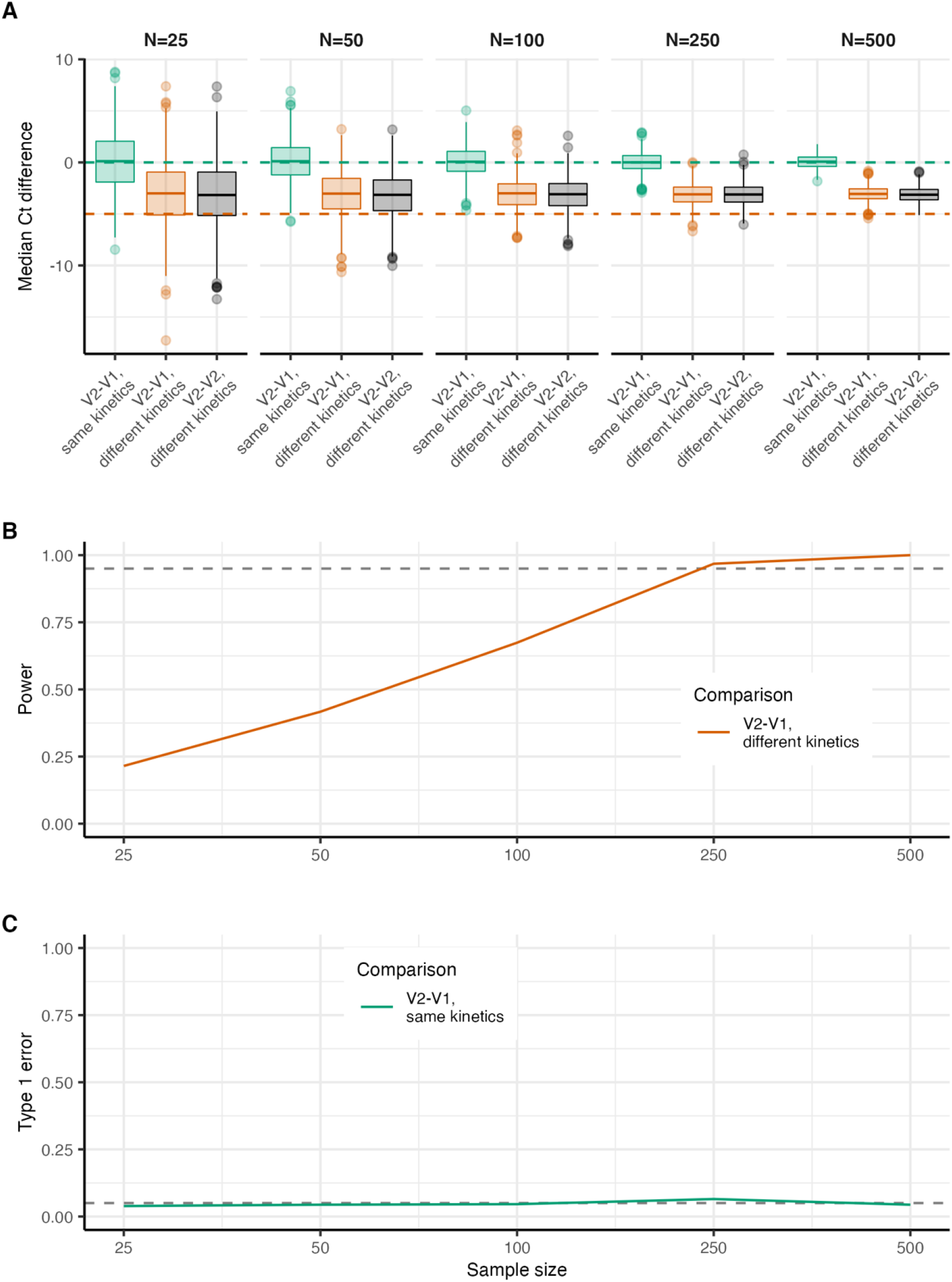
Accuracy and power when comparing Ct values from variants obtained through random cross-sectional surveillance when samples are obtained when both variants have a growth rate of 0.03 using the simulation shown in Figure 1. All statistical tests are two-sided Wilcoxon rank sum tests. (**A**) Boxplots show the interquartile range (IQR, 75th to 25th percentile) and median across 1000 simulations for the difference in Ct value when comparing the original variant (V1), the new variant (V2) with identical kinetics, or the new variant (V2) with different kinetics. Whiskers show 1.5 times the largest and smallest values within 1.5 times the IQR. Dots show individual simulations outside 1.5 times the IQR. Horizontal dashed green lines show the true difference in median Ct values between the original variant and the new variant with identical kinetics. Horizontal dashed orange line shows the difference in peak Ct value assumed for the new variant with different kinetics relative to the original variant. For an entirely accurate test, the green dots should all align on the green horizontal line, and the orange and black dots should be identical and negative. (**B**) Assessment of empirical power to detect a true difference in Ct values at different sample sizes. (**C**) Probability of type 1 error (incorrectly infer a difference in Ct value) at different sample sizes.

**Figure S3.**
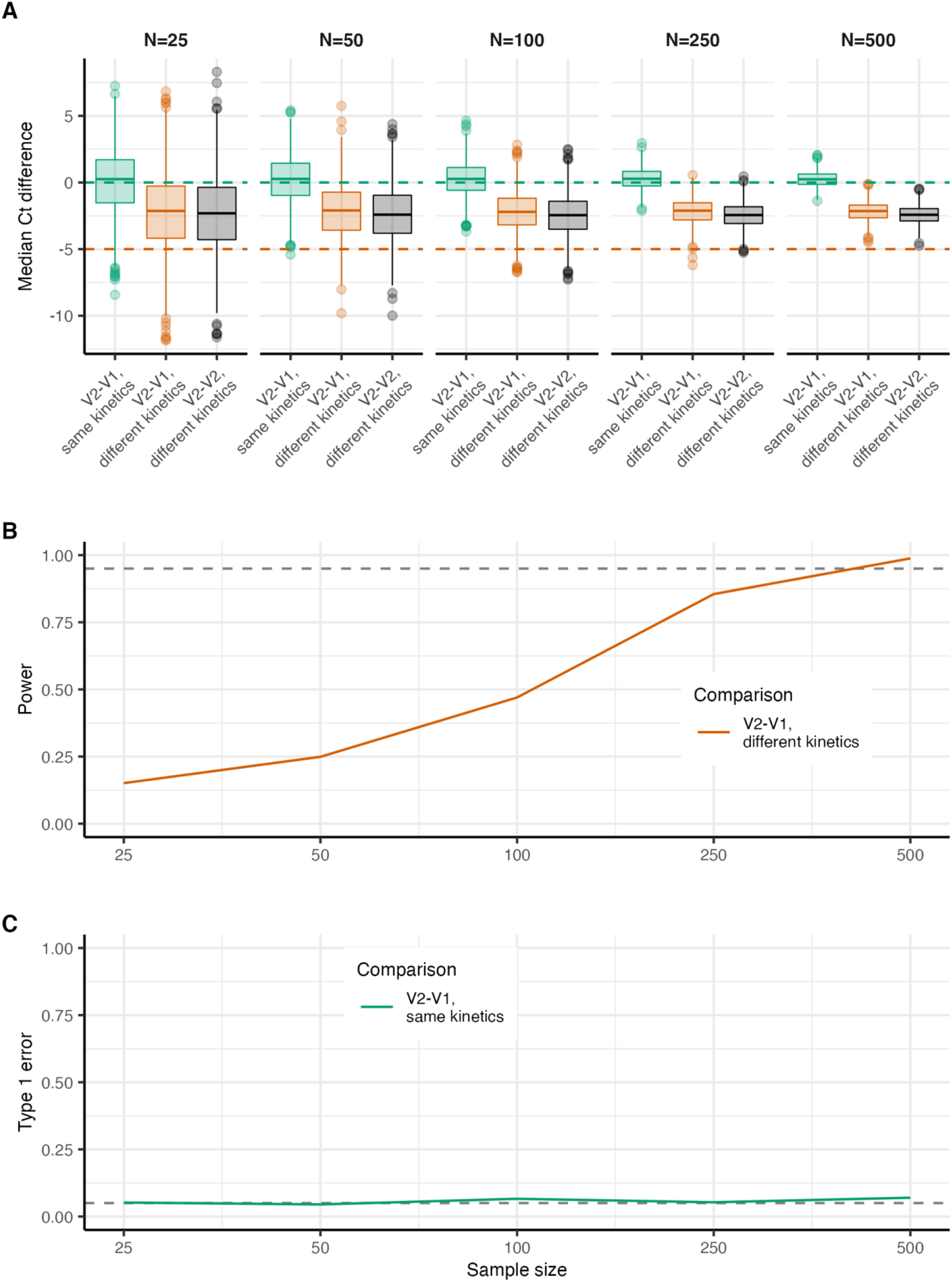
Accuracy and power when comparing Ct values from variants obtained through random cross-sectional surveillance when samples are obtained when both variants have a growth rate of -0.02 using the simulation shown in Figure 1. All statistical tests are two-sided Wilcoxon rank sum tests. (**A**) Boxplots show the interquartile range (IQR, 75th to 25th percentile) and median across 1000 simulations for the difference in Ct value when comparing the original variant (V1), the new variant (V2) with identical kinetics, or the new variant (V2) with different kinetics. Whiskers show 1.5 times the largest and smallest values within 1.5 times the IQR. Dots show individual simulations outside 1.5 times the IQR. Horizontal dashed green lines show the true difference in median Ct values between the original variant and the new variant with identical kinetics. Horizontal dashed orange line shows the difference in peak Ct value assumed for the new variant with different kinetics relative to the original variant. For an entirely accurate test, the green dots should all align on the green horizontal line, and the orange and black dots should be identical and negative. (**B**) Assessment of empirical power to detect a true difference in Ct values at different sample sizes. (**C**) Probability of type 1 error (incorrectly infer a difference in Ct value) at different sample sizes.

**Figure S4.**
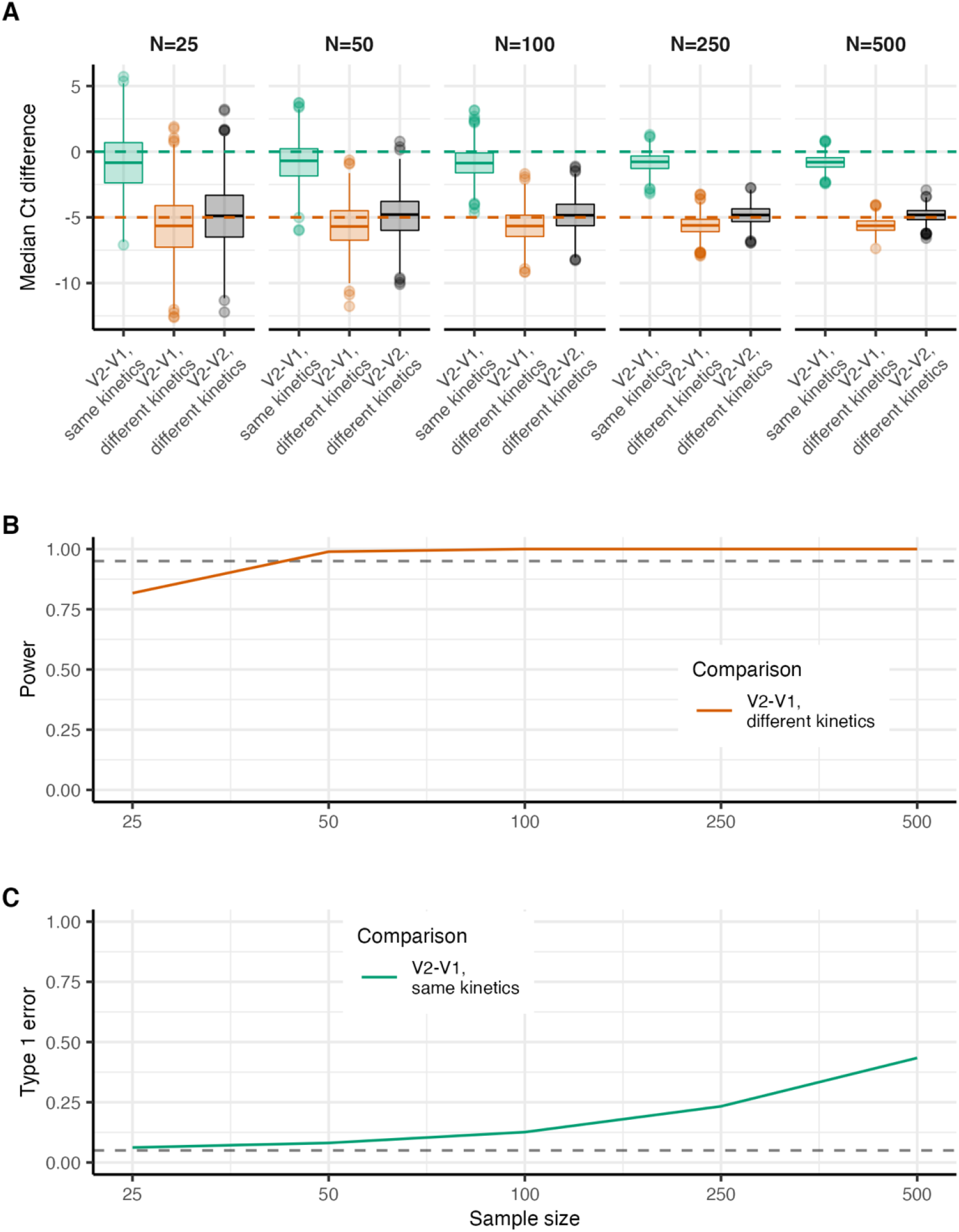
Accuracy and power when comparing Ct values from variants obtained through symptom-based surveillance when samples are obtained at day 270 of the simulation shown in Figure 3. All statistical tests are two-sided Wilcoxon rank sum tests. (**A**) Boxplots show the interquartile range (IQR, 75th to 25th percentile) and median across 1000 simulations for the difference in Ct value when comparing the original variant (V1), the new variant (V2) with identical kinetics, or the new variant (V2) with different kinetics. Whiskers show 1.5 times the largest and smallest values within 1.5 times the IQR. Dots show individual simulations outside 1.5 times the IQR. Horizontal dashed green lines show the true difference in median Ct values between the original variant and the new variant with identical kinetics. Horizontal dashed orange line shows the difference in peak Ct value assumed for the new variant with different kinetics relative to the original variant. For an entirely accurate test, the green dots should all align on the green horizontal line, and the orange and black dots should be identical and negative. (**B**) Assessment of empirical power to detect a true difference in Ct values at different sample sizes. (**C**) Probability of type 1 error (incorrectly infer a difference in Ct value) at different sample sizes.

**Figure S5.**
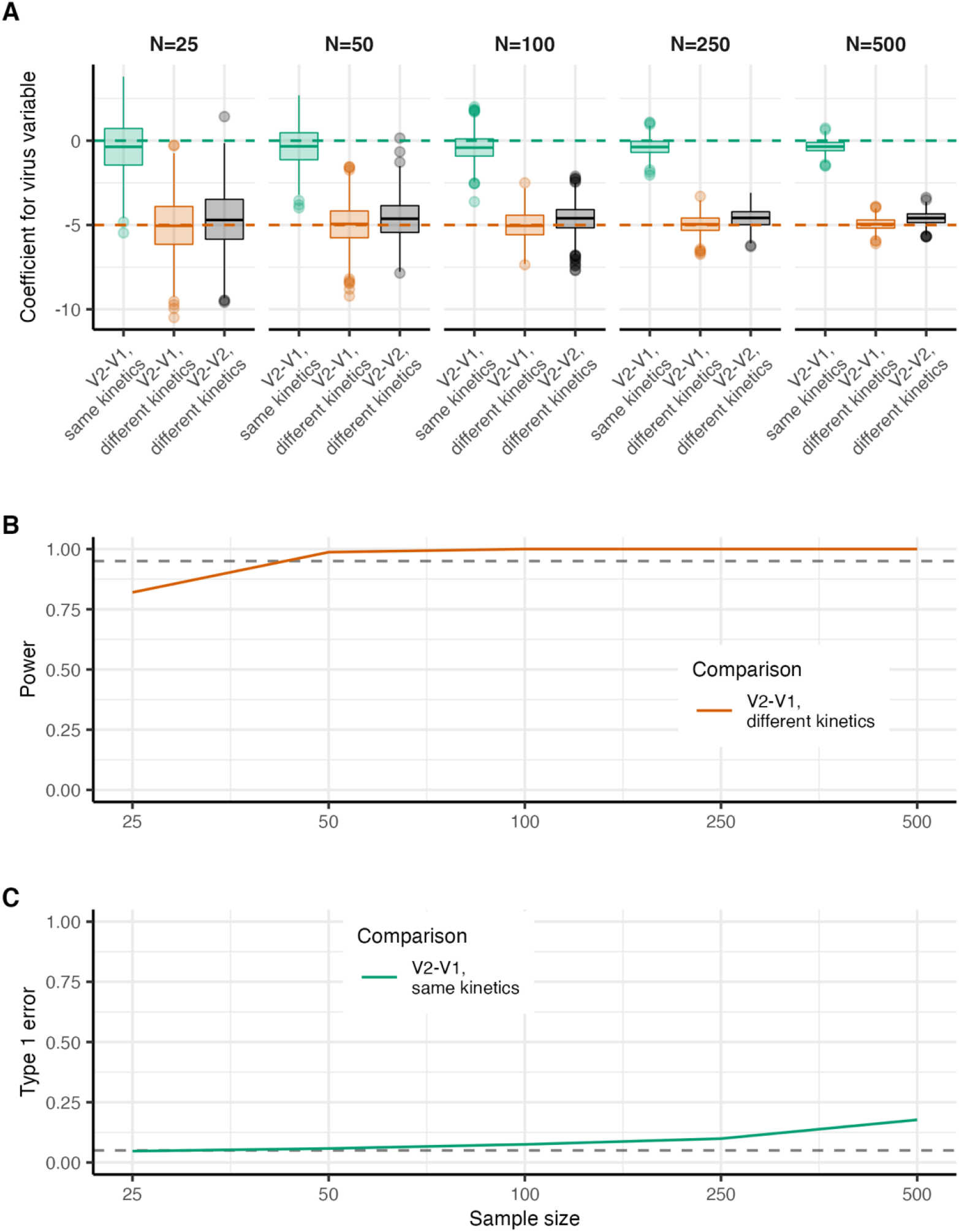
Accuracy and power when comparing Ct values from variants obtained through symptom-based surveillance when samples are obtained at day 270 of the simulation shown in Figure 3. All statistical tests are simple linear regression models as described in Materials and methods. (**A**) Boxplots show the interquartile range (IQR, 75th to 25th percentile) and median across 1000 simulations of the mean regression coefficient estimate for the effect of variant on difference in Ct value when comparing the original variant (V1), the new variant (V2) with identical kinetics, or the new variant (V2) with different kinetics. Whiskers show 1.5 times the largest and smallest values within 1.5 times the IQR. Dots show individual simulations outside 1.5 times the IQR. Horizontal dashed green lines show the true difference in median Ct values between the original variant and the new variant with identical kinetics. Horizontal dashed orange line shows the difference in peak Ct value assumed for the new variant with different kinetics relative to the original variant. For an entirely accurate test, the green dots should all align on the green horizontal line, and the orange and black dots should be identical and negative. (**B**) Assessment of empirical power to detect a true difference in Ct values at different sample sizes. (**C**) Probability of type 1 error (incorrectly infer a difference in Ct value) at different sample sizes.

**Figure S6.**
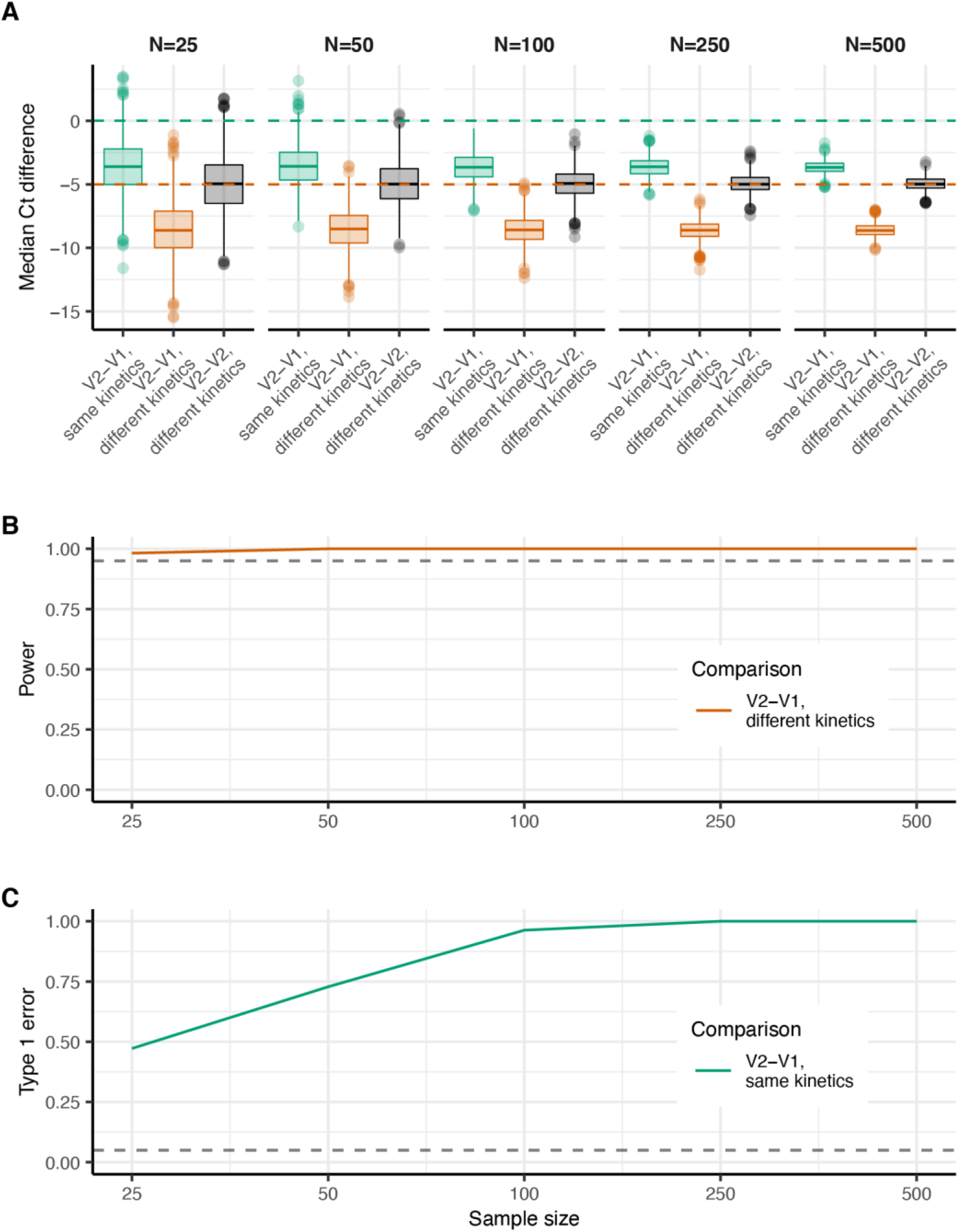
Identical to Figure S4, but assuming that the sampling delay distribution of original variant has a slightly higher mean and standard deviation than the new variant (shape parameter and shape parameters of 7 and 0.9 respectively vs. 5).

**Figure S7.**
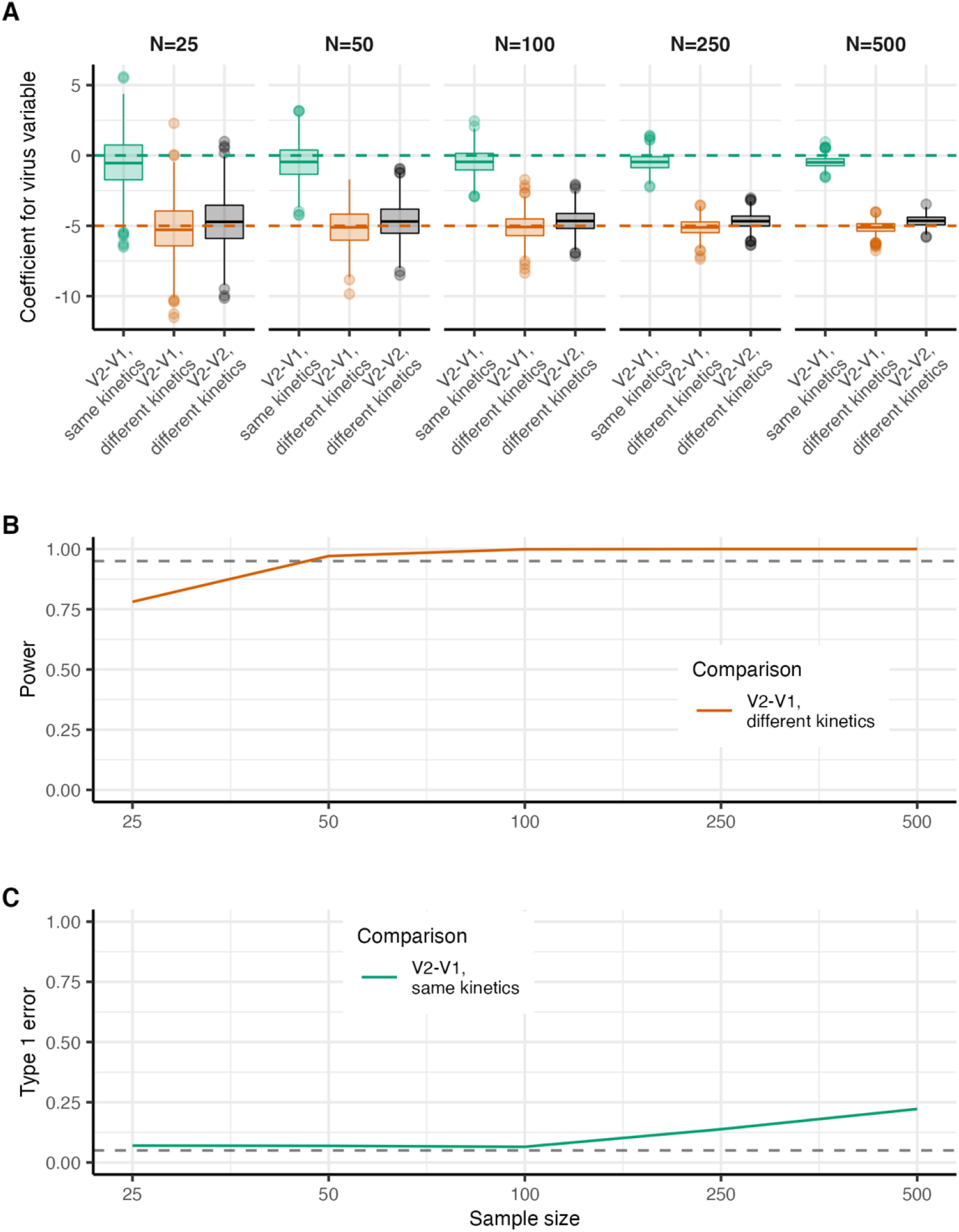
Identical to Figure S5, but assuming that the sampling delay distribution of original variant has a slightly higher mean and standard deviation than the new variant (shape parameter and shape parameters of 7 and 0.9 respectively vs. 5).

**Figure S8.**
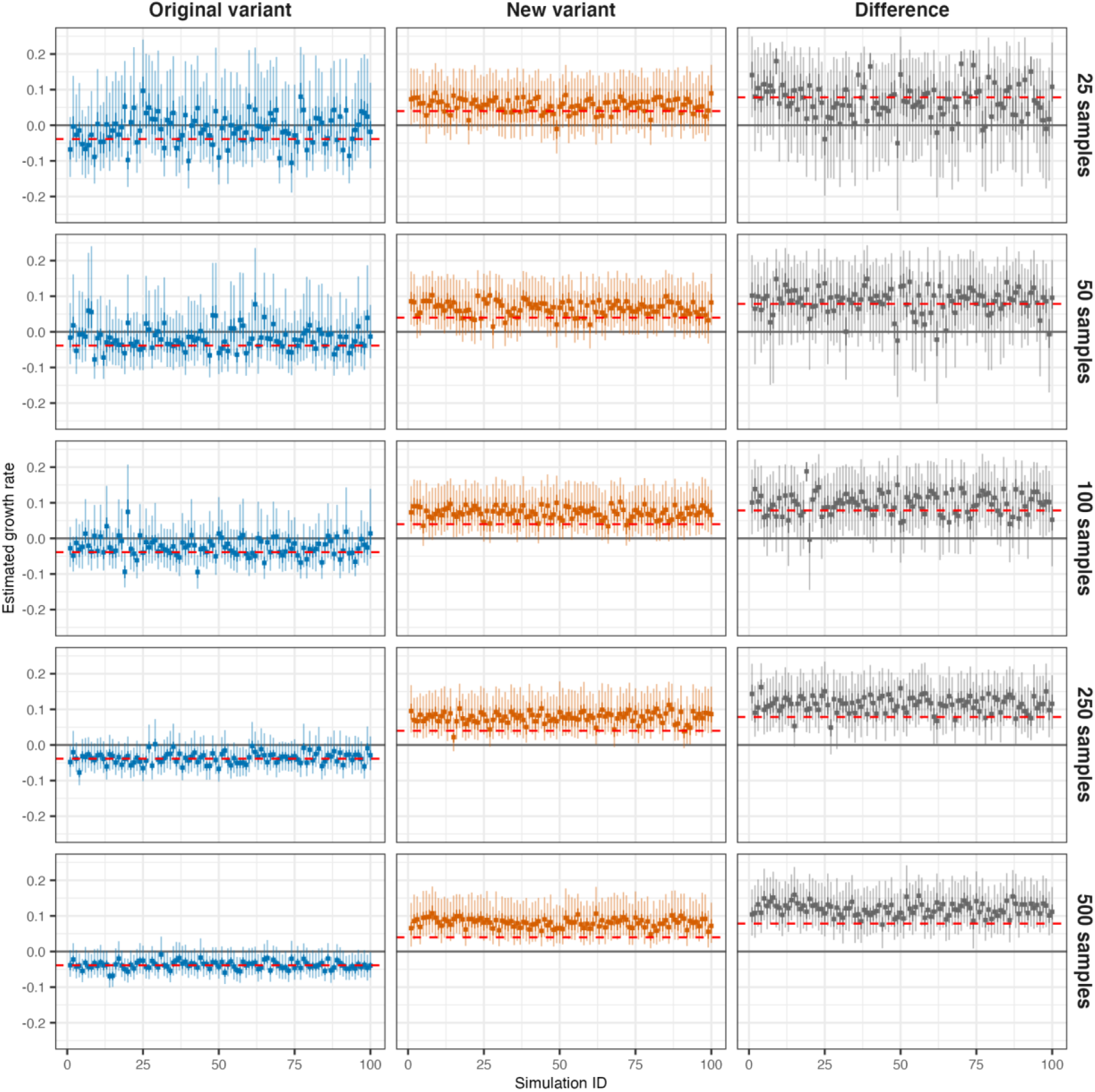
Estimated growth rates and variant differences compared to true simulated values, assuming that samples are obtained at day 270 of the simulation shown in Figure 1. The original variant is in epidemic decline, whereas the new variant is in epidemic growth. Rows show increasing sample sizes. Each point-range plot shows the posterior mean (dot), 50% CrI (dark line) and 95% CrI (faint line) for a distinct simulation. Dashed red line shows true value in the simulation.

**Figure S9.**
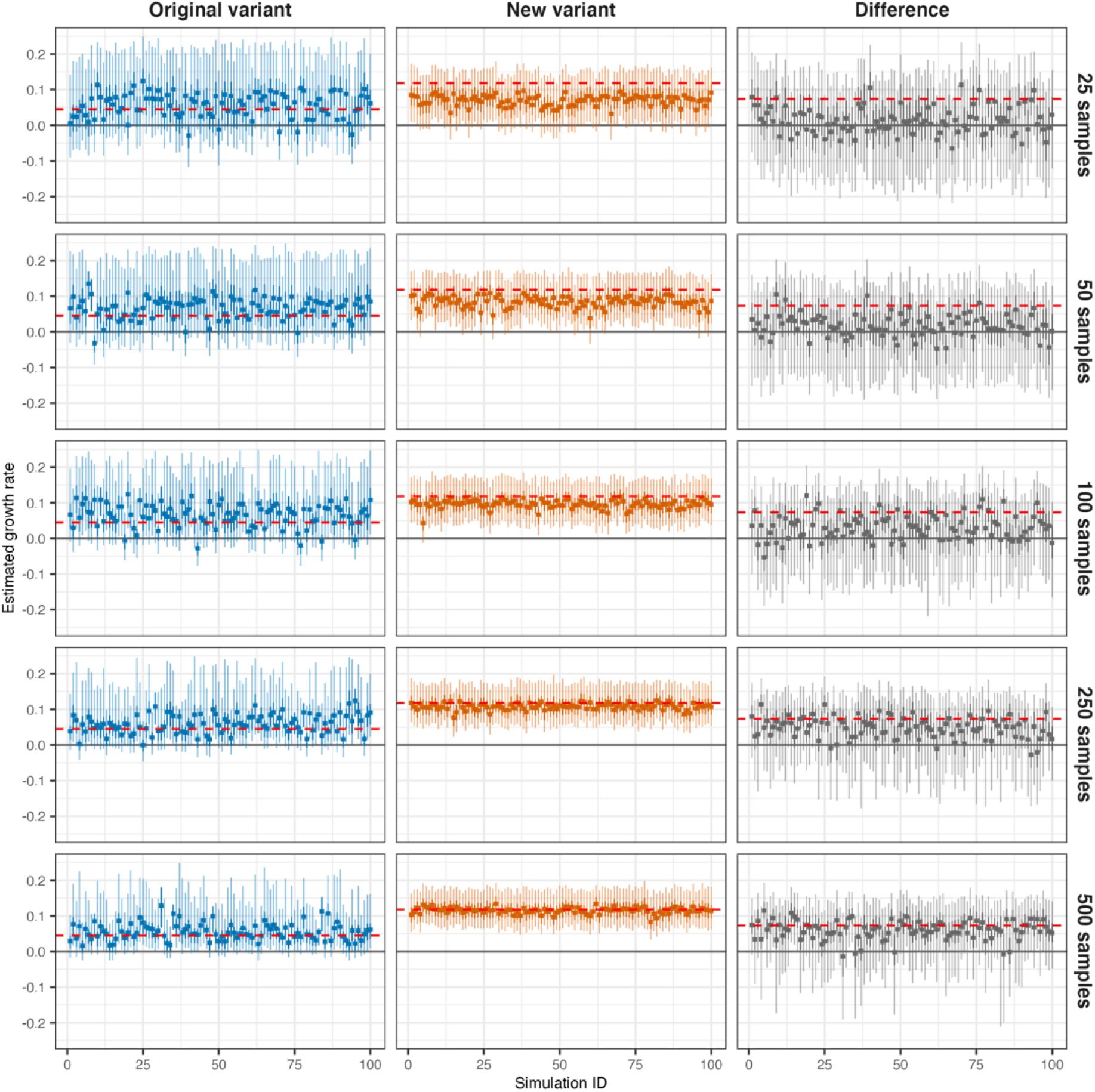
Estimated growth rates and variant differences compared to true simulated values, assuming that samples are obtained from a simulation where both variants are increasing simultaneously. The original variant has R_0_=1.5, whereas the new variant has R_0_=2.5. Rows show increasing sample sizes. Each point-range plot shows the posterior mean (dot), 50% CrI (dark line) and 95% CrI (faint line) for a distinct simulation. Dashed red line shows true value in the simulation.

**Figure S10.**
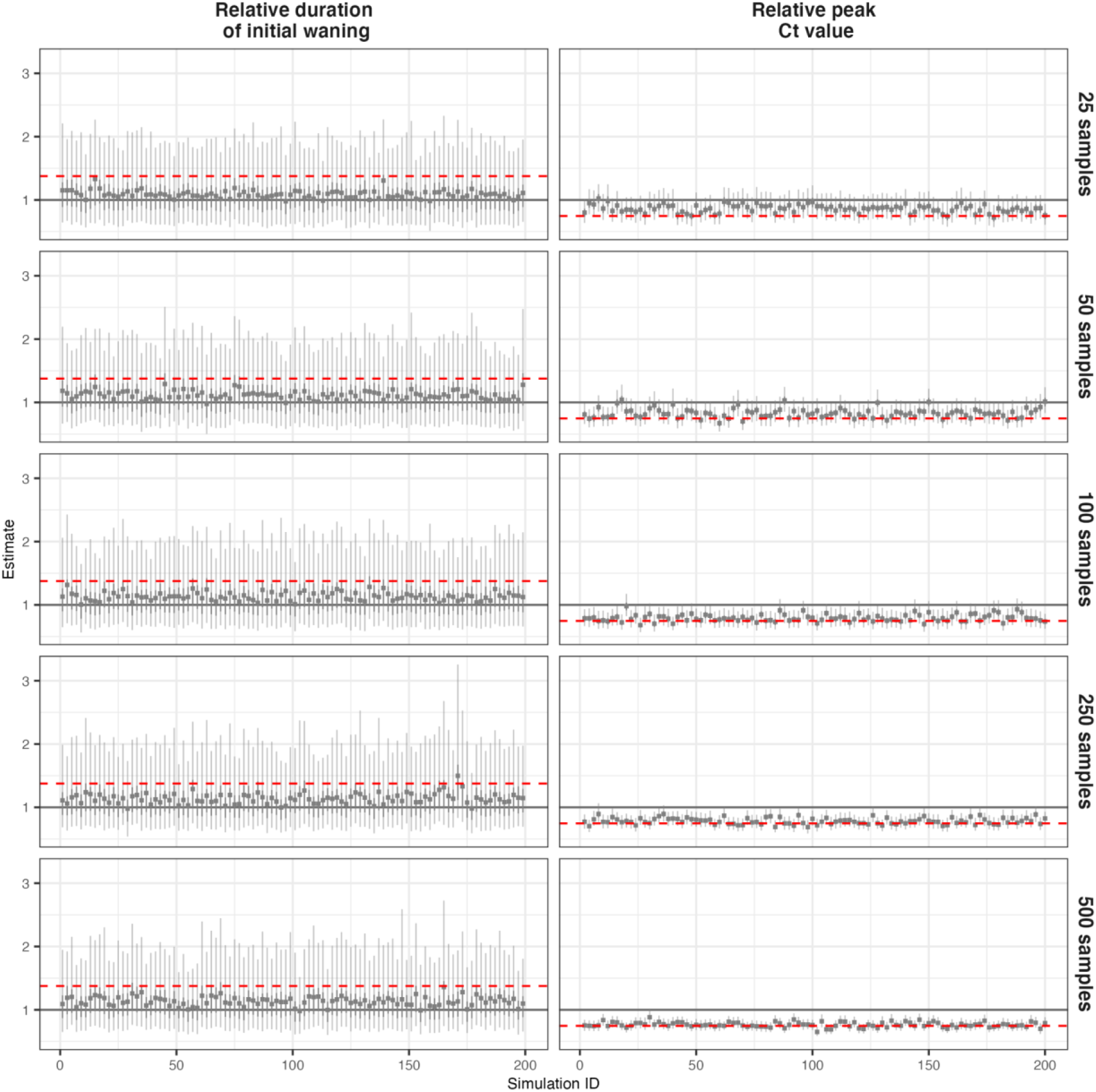
Posterior estimates for differences in viral kinetics between the two variants, assuming that samples are obtained at day 270 of the simulation shown in Figure 1. The original variant is in epidemic decline, whereas the new variant is in epidemic growth. Rows show increasing sample sizes. Each point-range plot shows the posterior mean (dot), 50% CrI (dark line) and 95% CrI (faint line) for a distinct simulation. Dashed red line shows true value in the simulation.

**Figure S11.**
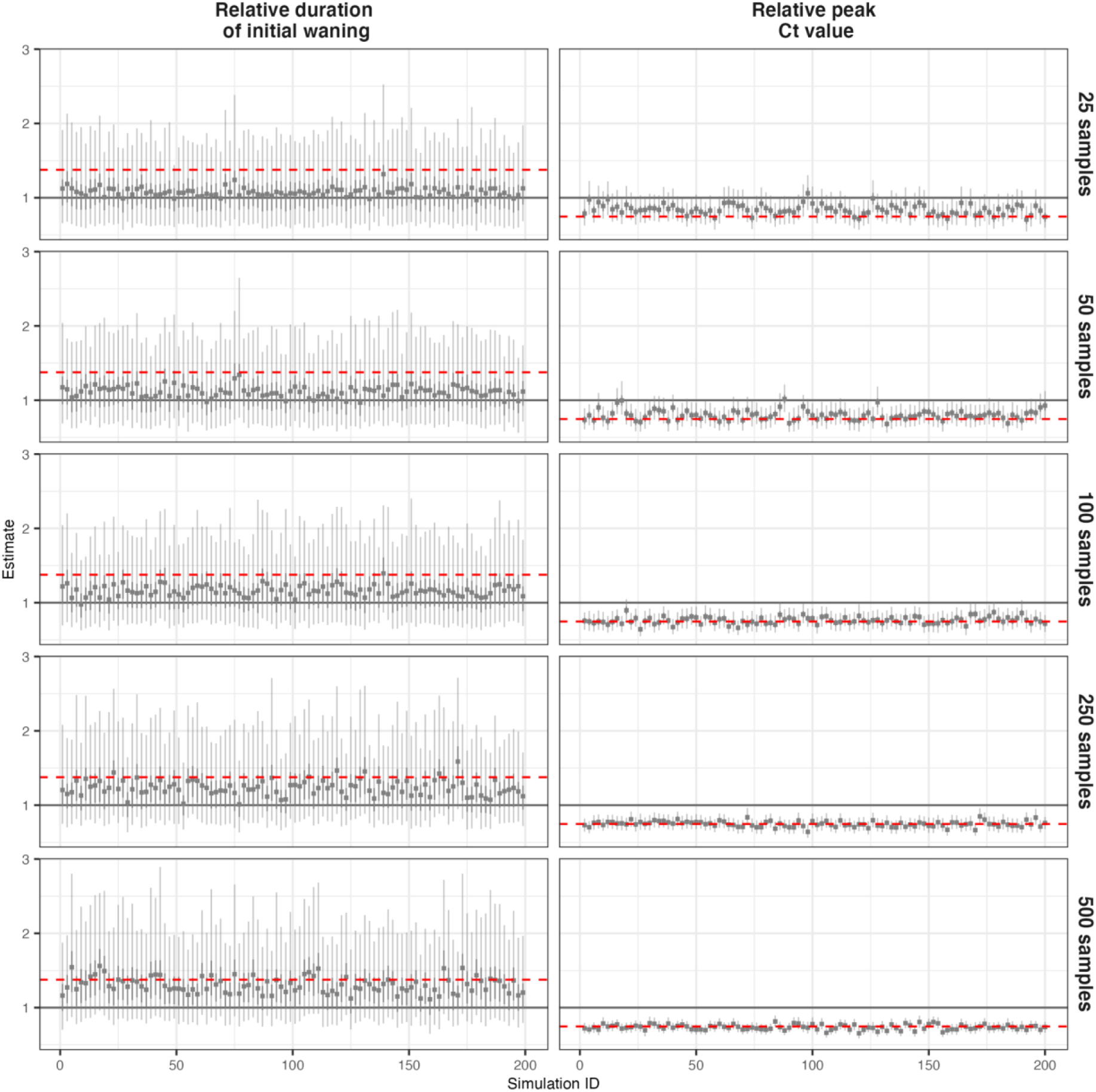
Posterior estimates for differences in viral kinetics between the two variants, assuming that samples are obtained at day 270 of the simulation shown in Figure 1. The original variant has R_0_=1.5, whereas the new variant has R_0_=2.5. Rows show increasing sample sizes. Each point-range plot shows the posterior mean (dot), 50% CrI (dark line) and 95% CrI (faint line) for a distinct simulation. Dashed red line shows true value in the simulation.

**Table S1:**
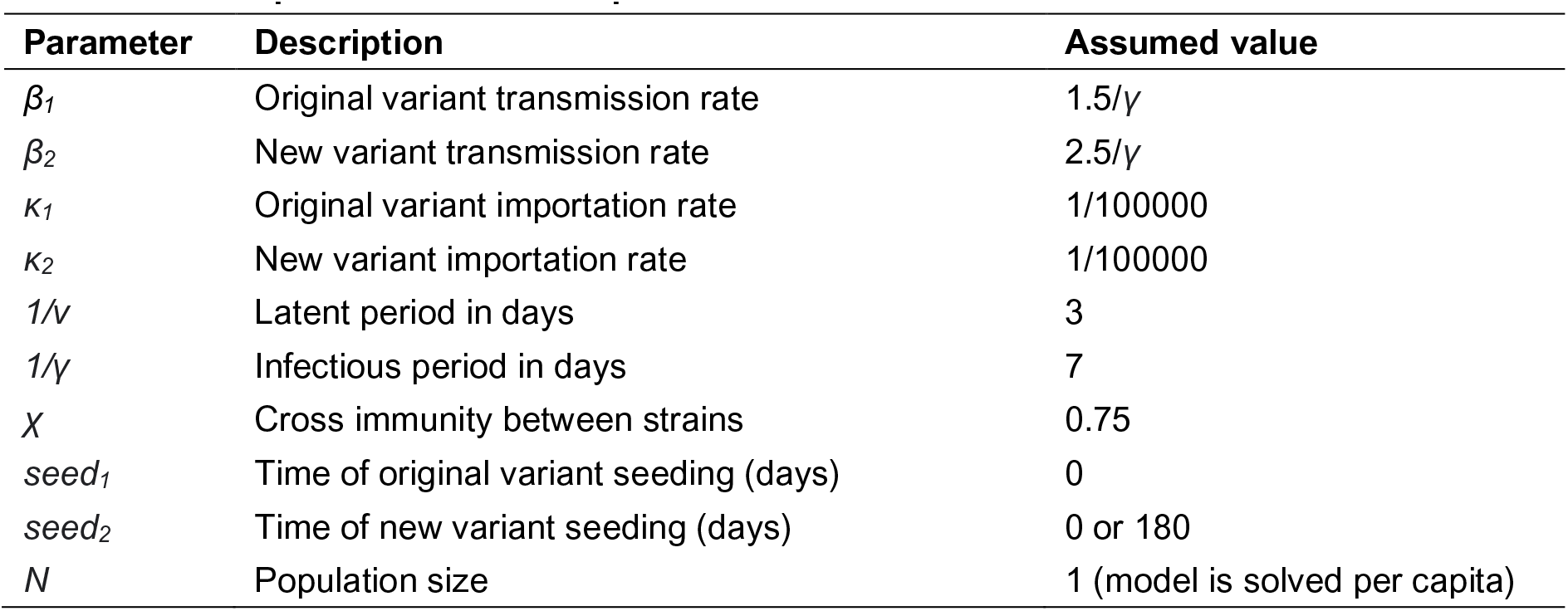
Description of SEIR model parameters

| Parameter | Description | Assumed value |
| --- | --- | --- |
| $\beta_1$ | Original variant transmission rate | $1.5/\gamma$ |
| $\beta_2$ | New variant transmission rate | $2.5/\gamma$ |
| $\kappa_1$ | Original variant importation rate | $1/100000$ |
| $\kappa_2$ | New variant importation rate | $1/100000$ |
| $1/\nu$ | Latent period in days | 3 |
| $1/\gamma$ | Infectious period in days | 7 |
| $\chi$ | Cross immunity between strains | 0.75 |
| $seed_1$ | Time of original variant seeding (days) | 0 |
| $seed_2$ | Time of new variant seeding (days) | 0 or 180 |
| $N$ | Population size | 1 (model is solved per capita) |

**Table S2:** Description of viral kinetics model parameters

| Parameter | Description | Point estimate | Prior |
| --- | --- | --- | --- |
| **Viral kinetics model** |  |  |  |
| $t_e$ | Time from infection to initial viral growth | 1.00 days | Fixed |
| $C_0$ | Ct value at time of infection | 45.0 | Fixed |
| $t_p$ | Days from initial viral growth to peak viral load | 5.00 days | Fixed |
| $C_p$ | Modal Ct value at peak viral load | 19.7 | Normal(19.7, 2.00) |
| $t_s$ | Time from peak viral load to plateau phase | 13.3 days | Normal(13.3, 3.00) |
| $C_s$ | Modal Ct value at $a = t_{eclipse} + t_{peak} + t_{switch}$ | 38.0 | Normal(38.0, 1.00) |
| $p_{addl}$ | Daily probability of detectability loss after $t_{switch}$ | 0.103 | Beta(10.5, 91.2) |
| $C_{LOD}$ | Limit of detection of Ct value | 40.0 | Fixed |
| $\sigma_{obs}$ | Initial scale parameter for the Gumbel distribution until $a = t_e + t_p + t_s$ | 5.00 | Normal(5.00, 0.50) |
| $s_{mod}$ | Multiplicative factor applied to scale parameter for the Gumbel distribution starting at $a = t_{eclipse} + t_{peak} + t_{switch} + t_{scale}$ | 0.789 | Fixed |
| $t_{mod}$ | Time from secondary waning phase until Gumbel distribution reaches its minimum scale parameter | 14.0 days | Fixed |
| **New variant viral kinetics** |  |  |  |
| $\rho$ | Peak Ct value of new variant relative to original variant | 0.747 (5 Ct values lower) | Log-normal(0, 0.5) (on linear scale) |
| $\eta$ | Time from peak to plateau phase for new variant relative to original variant | 1.38 (5 days longer) | Log-normal(0, 0.5) (on linear scale) |
| **Exponential growth model** |  |  |  |
| $\beta_1$ | Original variant exponential growth rate | NA | Normal(0, 0.1) |
| $\beta_2$ | New variant exponential growth rate | NA | Normal(0, 0.1) |

## Notes

### Author Declarations

Not applicable. All data used in this study were generated through computer simulations.

## References

1. Abdool Karim SS, de Oliveira T. New SARS-CoV-2 Variants - Clinical, Public Health, and Vaccine Implications. N Engl J Med. 2021;384: 1866–1868.

2. Gog JR, Grenfell BT. Dynamics and selection of many-strain pathogens. Proc Natl Acad Sci U S A. 2002;99: 17209–17214.

3. Bedford T, Riley S, Barr IG, Broor S, Chadha M, Cox NJ, et al. Global circulation patterns of seasonal influenza viruses vary with antigenic drift. Nature. 2015;523: 217– 220.

4. Russell CA, Jones TC, Barr IG, Cox NJ, Garten RJ, Gregory V, et al. The global circulation of seasonal influenza A (H3N2) viruses. Science. 2008;320: 340–346.

5. Faria NR, Mellan TA, Whittaker C, Claro IM, Candido D da S, Mishra S, et al. Genomics and epidemiology of the P.1 SARS-CoV-2 lineage in Manaus, Brazil. Science. 2021;372: 815–821.

6. Davies NG, Jarvis CI, CMMID COVID-19 Working Group, Edmunds WJ, Jewell NP, Diaz-Ordaz K, et al. Increased mortality in community-tested cases of SARS-CoV-2 lineage B.1.1.7. Nature. 2021;593: 270–274.

7. Threat Assessment Brief: Implications for the EU/EEA on the spread of the SARS-CoV-2 Delta (B.1.617.2) variant of concern. 23 Jun 2021 [cited 13 Jul 2021]. Available: https://www.ecdc.europa.eu/en/publications-data/threat-assessment-emergence-and-impact-sars-cov-2-delta-variant

8. . [No title]. [cited 13 Jul 2021]. Available: https://assets.publishing.service.gov.uk/government/uploads/system/uploads/attachment_data/file/1001358/Variants_of_Concern_VOC_Technical_Briefing_18.pdf

9. Volz E, Mishra S, Chand M, Barrett JC, Johnson R, Geidelberg L, et al. Assessing transmissibility of SARS-CoV-2 lineage B.1.1.7 in England. Nature. 2021;593: 266–269.

10. Tegally H, Wilkinson E, Giovanetti M, Iranzadeh A, Fonseca V, Giandhari J, et al. Emergence and rapid spread of a new severe acute respiratory syndrome-related coronavirus 2 (SARS-CoV-2) lineage with multiple spike mutations in South Africa. medRxiv. 2020; 2020.12.21.20248640.

11. Dhar MS, Marwal R, Radhakrishnan VS, Ponnusamy K, Jolly B, Bhoyar RC, et al. Genomic characterization and Epidemiology of an emerging SARS-CoV-2 variant in Delhi, India. medRxiv. 2021; 2021.06.02.21258076.

12. Morris DH, Gostic KM, Pompei S, Bedford T, Łuksza M, Neher RA, et al. Predictive Modeling of Influenza Shows the Promise of Applied Evolutionary Biology. Trends Microbiol. 2018;26: 102–118.

13. Harvey WT, Carabelli AM, Jackson B, Gupta RK, Thomson EC, Harrison EM, et al. SARS-CoV-2 variants, spike mutations and immune escape. Nat Rev Microbiol. 2021;19: 409–424.

14. MacLean OA, Orton RJ, Singer JB, Robertson DL. No evidence for distinct types in the evolution of SARS-CoV-2. Virus Evol. 2020;6. doi:10.1093/ve/veaa034

15. Cai Y, Zhang J, Xiao T, Lavine CL, Rawson S, Peng H, et al. Structural basis for enhanced infectivity and immune evasion of SARS-CoV-2 variants. Science. 2021. doi:10.1126/science.abi9745

16. Jones TC, Biele G, Mühlemann B, Veith T, Schneider J, Beheim-Schwarzbach J, et al. Estimating infectiousness throughout SARS-CoV-2 infection course. Science. 2021;373. doi:10.1126/science.abi5273

17. Lee LYW, Rozmanowski S, Pang M, Charlett A, Anderson C, Hughes GJ, et al. Severe Acute Respiratory Syndrome Coronavirus 2 (SARS-CoV-2) Infectivity by Viral Load, S Gene Variants and Demographic Factors, and the Utility of Lateral Flow Devices to Prevent Transmission. Clin Infect Dis. 2021 [cited 14 Jul 2021]. doi:10.1093/cid/ciab421

18. Marks M, Millat-Martinez P, Ouchi D, et al. Transmission of COVID-19 in 282 clusters in Catalonia, Spain: a cohort study. Lancet Infect Dis 2021; 5: 629–36. https://doi.org/10.1016/S1473-3099(21)00398-4.

19. Bjorkman KK, Saldi TK, Lasda E, Bauer LC, Kovarik J, Gonzales PK, Fink MR, Tat KL, Hager CR, Davis JC, Ozeroff CD, Brisson GR, Larremore DB, Leinwand LA, McQueen MB, Parker R. Higher viral load drives infrequent SARS-CoV-2 transmission between asymptomatic residence hall roommates. J Infect Dis. 2021. doi:10.1093/infdis/jiab386.

20. Kissler SM, Fauver JR, Mack C, Tai CG, Breban MI, Watkins AE, et al. Densely sampled viral trajectories for SARS-CoV-2 variants alpha (B.1.1.7) and epsilon (B.1.429). medRxiv. 2021; 2021.02.16.21251535.

21. Calistri P, Amato L, Puglia I, Cito F, Di Giuseppe A, Danzetta ML, et al. Infection sustained by lineage B.1.1.7 of SARS-CoV-2 is characterised by longer persistence and higher viral RNA loads in nasopharyngeal swabs. Int J Infect Dis. 2021;105: 753–755.

22. Müller NF, Wagner C, Frazar CD, Roychoudhury P, Lee J, Moncla LH, et al. Viral genomes reveal patterns of the SARS-CoV-2 outbreak in Washington State. Sci Transl Med. 2021;13. doi:10.1126/scitranslmed.abf0202

23. Roquebert B, Haim-Boukobza S, Trombert-Paolantoni S, Lecorche E, Verdurme L, Foulongne V, et al. SARS-CoV-2 variants of concern are associated with lower RT-PCR amplification cycles between January and March 2021 in France. medRxiv. doi:10.1101/2021.03.19.21253971

24. Borges V, Sousa C, Menezes L, Gonçalves AM, Picão M, Almeida JP, et al. Tracking SARS-CoV-2 lineage B.1.1.7 dissemination: insights from nationwide spike gene target failure (SGTF) and spike gene late detection (SGTL) data, Portugal, week 49 2020 to week 3 2021. Eurosurveillance. 2021. doi:10.2807/1560-7917.es.2021.26.10.2100130

25. Frampton D, Rampling T, Cross A, Bailey H, Heaney J, Byott M, et al. Genomic characteristics and clinical effect of the emergent SARS-CoV-2 B.1.1.7 lineage in London, UK: a whole-genome sequencing and hospital-based cohort study. The Lancet Infectious Diseases. 2021. doi:10.1016/s1473-3099(21)00170-5

26. Kidd M, Richter A, Best A, Cumley N, Mirza J, Percival B, et al. S-Variant SARS-CoV-2 Lineage B1.1.7 Is Associated With Significantly Higher Viral Load in Samples Tested by TaqPath Polymerase Chain Reaction. J Infect Dis. 2021;223: 1666–1670.

27. Hay JA, Kennedy-Shaffer L, Kanjilal S, Lennon NJ, Gabriel SB, Lipsitch M, et al. Estimating epidemiologic dynamics from cross-sectional viral load distributions. Science. 2021. doi:eabh0635.

28. Walker AS, Pritchard E, House T, Robotham JV, Birrell PJ, Bell I, et al. Ct threshold values, a proxy for viral load in community SARS-CoV-2 cases, demonstrate wide variation across populations and over time. Elife. 2021;10. doi:10.7554/eLife.64683

29. Alizon S, Selinger C, Sofonea MT, Haim-Boukobza S, Giannoli J-M, Ninove L, et al. Epidemiological and clinical insights from SARS-CoV-2 RT-PCR cycle amplification values. medRxiv. 2021; 2021.03.15.21253653.

30. Gostic KM, Kucharski AJ, Lloyd-Smith JO. Effectiveness of traveller screening for emerging pathogens is shaped by epidemiology and natural history of infection. Elife. 2015;4. doi:10.7554/eLife.05564

31. Grubaugh ND, Hanage WP, Rasmussen AL. Making Sense of Mutation: What D614G Means for the COVID-19 Pandemic Remains Unclear. Cell. 2020. pp. 794–795.

32. SARS-CoV-2, SARS-CoV, and MERS-CoV viral load dynamics, duration of viral shedding, and infectiousness: a systematic review and meta-analysis. The Lancet Microbe. 2021;2: e13–e22.

33. Eales O, Page AJ, Tang SN, Walters CE, Wang H, Haw D, et al. SARS-CoV-2 lineage dynamics in England from January to March 2021 inferred from representative community samples. doi:10.1101/2021.05.08.21256867

34. Golubchik T, Lythgoe KA, Hall M, Ferretti L, Fryer HR, MacIntyre-Cockett G, et al. Early analysis of a potential link between viral load and the N501Y mutation in the SARS-COV-2 spike protein. doi:10.1101/2021.01.12.20249080

35. Smith RL, Gibson LL, Martinez PP, Ke R, Mirza A, Conte M, et al. Longitudinal assessment of diagnostic test performance over the course of acute SARS-CoV-2 infection. J Infect Dis. 2021 [cited 16 Jul 2021]. doi:10.1093/infdis/jiab337

36. Li B, Deng A, Li K, Hu Y, Li Z, Xiong Q, et al. Viral infection and transmission in a large well-traced outbreak caused by the Delta SARS-CoV-2 variant. doi:10.1101/2021.07.07.21260122

37. Rhoads D, Peaper DR, She RC, Nolte FS, Wojewoda CM, Anderson NW, et al. College of American Pathologists (CAP) Microbiology Committee Perspective: Caution Must Be Used in Interpreting the Cycle Threshold (Ct) Value. Clinical infectious diseases: an official publication of the Infectious Diseases Society of America. 2021. pp. e685–e686.

38. Cendejas-Bueno E, Romero-Gómez MP, Escosa-García L, Jiménez-Rodríguez S, Mingorance J, García-Rodríguez J, et al. Lower nasopharyngeal viral loads in pediatric population. The missing piece to understand SARS-CoV-2 infection in children? J Infect. 2021. doi:10.1016/j.jinf.2021.06.009

39. Borremans B, Gamble A, Prager KC, Helman SK, McClain AM, Cox C, et al. Quantifying antibody kinetics and RNA detection during early-phase SARS-CoV-2 infection by time since symptom onset. Elife. 2020;9. doi:10.7554/eLife.60122

40. Kissler SM, Fauver JR, Mack C, Olesen SW, Tai C, Shiue KY, et al. Viral dynamics of acute SARS-CoV-2 infection and applications to diagnostic and public health strategies. PLOS Biology. 2021. p. e3001333. doi:10.1371/journal.pbio.3001333

41. Riley S, Wang H, Eales O, Haw D, Walters CE, Ainslie KEC, et al. REACT-1 round 12 report: resurgence of SARS-CoV-2 infections in England associated with increased frequency of the Delta variant. medRxiv. 2021; 2021.06.17.21259103.

42. Decreased infectivity following BNT162b2 vaccination: A prospective cohort study in Israel. The Lancet Regional Health - Europe. 2021;7: 100150.

43. Brown KA, Gubbay J, Hopkins J, Patel S, Buchan SA, Daneman N, et al. S-Gene Target Failure as a Marker of Variant B.1.1.7 Among SARS-CoV-2 Isolates in the Greater Toronto Area, December 2020 to March 2021. JAMA. 2021. p. 2115. doi:10.1001/jama.2021.5607

44. Kissler SM, Tedijanto C, Goldstein E, Grad YH, Lipsitch M. Projecting the transmission dynamics of SARS-CoV-2 through the postpandemic period. Science. 2020;368: 860–868.

45. J. A. Hay, L. Kennedy-Shaffer, jameshay218/virosolver: Publication release, version v1.0.2, Zenodo (2021); http://doi.org/10.5281/zenodo.4776812.

46. S. A. Lauer, K. H. Grantz, Q. Bi, F. K. Jones, Q. Zheng, H. R. Meredith, A. S. Azman, N. G. Reich, J. Lessler. The incubation period of coronavirus disease 2019 (COVID-19) from publicly reported confirmed cases: estimation and application. Ann. Int. Med. 172, 577–582 (2020).

